# PAGAN predicts digenic interactions by generalizing single-gene representations in biological networks

**DOI:** 10.64898/2026.01.27.26344931

**Authors:** Romain Nicolle, Barthélémy Caron, Valérie Malan, Antonio Rausell

## Abstract

Digenic alterations can produce phenotypes such as synthetic lethality or digenic disease that are not observed upon individual gene perturbation, often by disrupting compensatory or redundant biological mechanisms. We hypothesized that gene pairs underlying such phenotypes share, when considered jointly, biological network properties analogous to those of essential genes or monogenic Mendelian disease genes. To test this hypothesis, we developed PAGAN, a graph representation learning framework that learns phenotypically relevant network signatures from single-gene labels in heterogeneous biological knowledge graphs and generalizes them to gene pairs without explicit pairwise supervision. PAGAN represents each gene pair as a new node embedded in the same network context as individual genes, enabling inference of pair-level properties from their combined neighborhood. Using multiplex knowledge-based networks in yeast and human, we show that PAGAN predicts synthetic lethal interactions and digenic disease gene pairs by training only on essential genes or monogenic disease genes, respectively. Across multiple evaluation settings, PAGAN achieves competitive performance relative to supervised state-of-the-art methods while avoiding reliance on currently limited and biased catalogs of known gene pairs. This framework provides a scalable strategy to explore combinatorial genetic architectures and prioritize candidate digenic interactions in functional genomics and rare disease diagnostics.

## Introduction

Current diagnostic rates for rare diseases are heterogeneous across disease categories, reflecting differences in genetic architecture and available molecular knowledge (1). Despite the routine use of whole-genome sequencing (WGS), diagnostic yields typically reach on average 30%, leaving a large fraction of cases without a genetic diagnosis (2). This limitation is particularly evident in neurodevelopmental disorders (NDDs), which are characterized by marked clinical and genetic heterogeneity. Interpretation is challenged by the large number of genomic variants present in each individual, including approximately 5–10 million single-nucleotide variants (SNVs) and more than 20,000 structural variants (SVs) (3,4).

A working hypothesis is that a fraction of unresolved cases may involve more complex genetic architectures that are not adequately captured by monogenic models, including digenic modes of inheritance. Systematic studies in yeasts and recent works in humans have highlighted the potential of digenic alterations to lead to phenotypic consequences, i.e. synthetic lethality or digenic disease, that would not have otherwise been produced upon individual gene alteration. In such cases, the joint presence of genetic alterations affecting two genes may lead to phenotypic consequences that would not be observed upon alteration of each gene individually. These effects may arise from the disruption of compensatory or redundant cellular or physiological mechanisms, involving parallel or complementary molecular pathways or biological functions, thereby complicating variant interpretation under standard diagnostic frameworks.

In humans, one of the earliest examples of digenic inheritance identified is retinitis pigmentosa type 7, which results from pathogenic variants affecting both *ROM1* and *PRPH2* genes in a double-heterozygous configuration (5). Since this initial report, numerous cases have been described in which the observed phenotype was suspected or concluded to arise from the combined effect of genetic variations in two or more genes. These cases have been systematically collected and curated in the Digenic Diseases Database (DIDA), later expanded into the OLIgogenic Diseases Database (OLIDA) (6,7). The OLIDA database has enabled the development and evaluation of computational methods aimed at predicting gene pairs that may lead to digenic disease when jointly altered.

Current state-of-the-art approaches, including DiGePred, DIEP, VarCoPP2.0, and ARBOCK (8–11), rely on supervised learning frameworks trained on curated sets of gene pairs reported to be associated with human digenic diseases. Recently, Ogloblinsky et al. (12) conducted a systematic benchmarking of these methods using a reference dataset derived from OLIDA. In this benchmark, positive examples consisted of variant pairs involving two genes jointly associated with genetic disease, whereas negative examples comprised variant pairs involving two genes not implicated, as a pair, in genetic disorders. While this benchmark represents a valuable resource for method development and evaluation, the predictive performance of supervised approaches remains constrained by the limited number of currently known disease-associated gene pairs, which include only a few hundred gene pairs. Moreover, a large proportion of the reported disease-associated pairs are composed of either two monogenic Mendelian disease genes (MMDGs) or include at least one MMDG, resulting in a small set of recurrent genes across pairs. These characteristics limit the effective diversity of training data and pose challenges for supervised learning approaches that work directly on gene pairs.

Network-based analyses of biological systems, including protein-protein interaction, gene co-expression, and regulatory networks, have been widely used to identify candidate disease genes through computational approaches. A recent review by Sezin Kircali Ata *et al* (13) categorized existing network-based methods for disease gene prediction into three main classes. The first class comprises network diffusion approaches, which propagate information from known disease-associated genes through biological networks using random-walk or related diffusion processes. A second class includes machine learning methods that rely on handcrafted graph-derived features, which are extracted from network topology and subsequently used as input to conventional predictive models. Finally, graph representation learning approaches aim to automatically learn latent representations of genes and diseases directly from network structure, using techniques such as graph embeddings, or Graph Neural Networks (GNNs), thereby avoiding the need for manual feature engineering.

In this work, we hypothesized that pairs of genes leading to digenic phenotypes, could present, when jointly considered, biological network characteristics similar to those genes susceptible to lead to the analogous phenotypes caused upon monogenic alteration, i.e. gene essentiality or monogenic mendelian disease, respectively. In addition, we hypothesized that a machine learning algorithm suitable for representation learning on graph-structured data, such Graph Neural Networks, could learn such network characteristics through a supervised-learning approach. To address such hypothesis, we constructed species-specific knowledge-based networks for yeast and human, respectively, aggregating heterogeneous biological relations among genes and proteins, including protein interactions, gene expression in various cell types, signaling pathways, functional annotations, as well as specific gene characteristics such as sequence conservation metrics. We then trained diverse Graph Neural Networks on the identification of essential and Monogenic Mendelian disease genes on such heterogeneous networks and evaluated whether such models would generalize to the identification of analogous phenotypically-relevant gene-pair alterations leading, respectively, to synthetic lethality and digenic disease. From a more methodological perspective, we thus proposed to infer pair-of-gene characteristics by training Graph Neural Network models to predict their analogous individual gene characteristics (i.e. node classification) and apply it to classify an independent set of pairs of genes, where such pairs are jointly considered as if they were additional new individual nodes in the knowledge graph network. Thus, PAGAN is not trained on labeled gene pairs, but instead learns gene-pair properties by generalizing from single-gene labels. We named this approach as PAGAN: “PAirs of Genes As geNes” approach.

## Methods

### Human knowledge-based biological network

#### Nodes

A human knowledge-based biological network was built in the form of a multiplex heterogeneous graph. This network integrates five distinct types of nodes and eighteen edge types representing diverse biological relations (**Supplementary Table 1**). The five node types in the network represent nuclear protein-coding genes, anatomical structures, and three categories of Gene Ontology terms (14,15).The first node type, hereafter referred as GENE, represents protein-coding genes (and/or the proteins they encode) as defined in the Ensembl database (release 111, dated January 11th, 2024, downloaded on April 17th, 2024; (16)). Ensembl database was accessed using either the BioMart web interface (https://www.ensembl.org/) or the Python package pybiomart (https://github.com/jrderuiter/pybiomart). Only genes located on autosomes or sex chromosomes were retained; genes located on the mitochondrial chromosome or on alternative contigs were excluded. The second node type, ANATOMY, corresponds to organs, tissues or cell types and was derived from the Uberon ontology (v2022-12-13)(17–20). The remaining three node types represent terms from the Gene Ontology (GO): Biological Process (BP), Cellular Component (CC), and Molecular Function (MF). These ontology terms and their relations were extracted from the Gene Ontology Resource (https://www.geneontology.org/; release 2022_11_03, 10.5281/zenodo.7301602, (14,15)).

#### Edges

The network’s edges define relationships between the previous node types and consist of eighteen distinct types. Of these, the Protein-Protein Interaction (PPI) and Paralogous edges are undirected; thus, each of these is represented by two directed edges in opposite directions, ensuring symmetry. All other edge types are directed and were augmented with reverse edges to enable bidirectional message-passing operations during graph-based computations. Protein-protein interaction (PPI) edges connect pairs of GENE nodes and were sourced from version 12.0 of the STRING database (https://string-db.org/). Only high-confidence physical interactions between human proteins were retained (STRING combined score higher than 0.7). Paralogous edges also connect GENE nodes and were extracted from Ensembl release 108, dated October 20th, 2022, and downloaded on November 7th, 2022.). Gene Ontology relationships were derived from the ‘gene2go.gz’ file provided by NCBI (https://ftp.ncbi.nlm.nih.gov/gene/DATA/gene2go.gz, release 2022_11_03, 10.5281/zenodo.7301602). This file enabled construction of three edge types: GENE-to-BP, GENE-to-CC, and GENE-to-MF. Each of these was paired with a corresponding reverse edge as previously indicated. In addition, GO ontology-internal relationships were represented by edges connecting BP terms, edges connecting CC terms, and edges connecting MF terms. Each of these was supplemented by a reverse edge. These hierarchical ontology relations were parsed from the go-basic.obo file provided by Gene Ontology. The version used was format-version 1.2, released on November 3rd, 2022, and accessible at https://release.geneontology.org/2022-11-03/ontology/. Gene expression edges were added between GENE and ANATOMY nodes. These were obtained from the Bgee database (https://www.bgee.org/, version 15.1, (19,20). The file used was Homo_sapiens_expr_advanced.tsv.gz, downloaded from https://www.bgee.org/ftp/bgee_v15_1/download/calls/expr_calls/. This file includes expression associations between genes and anatomical entities from the Uberon integrative multi-species anatomy ontology (17,18), representing either organs or tissues. Finally, relationships between anatomical (i.e. edges between ANATOMY nodes), along with their corresponding reverse edges, were derived from the Uberon ext.obo file, (format-version: 1.2, data-version: uberon/releases/2022-12-13/ext.owl, obtained from https://github.com/obophenotype/uberon/releases/tag/v2022-12-13).

#### Node features

To characterize the protein-coding genes used as nodes in the knowledge graph, we compiled several gene-level constraint and intolerance metrics. These metrics were integrated as numerical features for each GENE node, with harmonization of gene identifiers to ensure consistency across data sources. First, the gene constraint metrics called “Z-scores for missense and synonymous variants” from the gnomAD v2 dataset were downloaded from the UCSC Genome Browser (https://hgdownload.soe.ucsc.edu/gbdb/hg38/gnomAD/pLI/) (21,22). Values were extracted from the missenseByGene.bb files, downloaded on January 13, 2023. The s_het_ value, an approximation to selection against heterozygous protein truncating variants, was sourced from Weghorn et al., 2019 (23). The Gene Damage Index (GDI) was retrieved from Itan et al., 2015 (24), providing an estimate of a gene’s tolerance to mutations based on population variation data. Finally, the FUSIL (Full Spectrum of Intolerance to Loss-of-function) classification for each gene was retrieved from Cacheiro et al., (25), which categorizes genes based on their degree of loss-of-function intolerance. Downstream supervised learning models for gene essentiality and synthetic lethality prediction developed in this work included only Z-score missense, Z-score synonymous, s_het_ value and GDI as node features in the knowledge graph. Downstream supervised learning models for disease gene prediction and digenic disease gene pairs prediction developed in this work used the previous four features plus the hot-encoded FUSIL classification as node features in the knowledge graph.

For nodes that did not represent genes (i.e., those of types ANATOMY, BP, CC, or MF), numerical features were derived based on their connectivity within the graph. Specifically, we assigned features by one-hot encoding the degree of each node, following suggestions and best practices discussed in community threads from the PyTorch Geometric team, accessible at the following URLs: https://github.com/pyg-team/pytorch_geometric/discussions/4546; https://github.com/pyg-team/pytorch_geometric/discussions/4746; https://github.com/pyg-team/pytorch_geometric/discussions/3844. This encoding enabled to assign to such node types with a structured numerical feature vector, making them suitable for downstream training using graph neural network (GNN) architectures.

Other human gene features used in this work for characterization purposes but not as predictive features included the Gene’s Disease Specificity Index (DSI) and the Disease Pleiotropy Index (DPI) as provided by DISGENET database (version 25.3; (26)). Both metrics are aimed at indicating how specific is a gene with respect to the associated diseases. A value of the DSI close to one means that the gene is disease-specific, while a value close to zero indicates that the gene is disease-promiscuous. The Disease Pleiotropy Index considers if the diseases associated with the gene are similar among them and belong to the same disease class or belong to different disease classes. In this case, disease–promiscuous genes generate values of DPI close to one.

#### Gene labels and pair-of-genes labels: gene essentiality, synthetic lethality, monogenic and digenic disease genes

We obtained several datasets allowing to assign GENE node labels and pair-of-GENE-nodes labels to the human knowledge graph, which will constitute the target labels we will aim to predict along the different GNN predictive tasks in this work, i.e.: gene essentiality, synthetic lethality, monogenic and digenic disease genes.

The definition of “gene essentiality” and “non-essentiality” commonly found in the literature is the following: a gene is considered “essential” if its biallelic inactivation leads to cell death, whereas a gene is considered “non-essential” if its biallelic inactivation is tolerated by the cell (27). The “essentiality” of a gene can be assessed by CRISPR and RNAi experimental methods allowing to target specific genes of interest in order to prevent their expression. However, these *in vitro* results are often cell-lines dependent and cannot be easily generalized. Indeed, the cell lines used to assess essentiality *in vitro* often correspond to immortalized cancer cell-lines; as such, they carry genetic variations activating or deactivating compensatory pathways allowing the cell to survive in conditions that would have been otherwise lethal, which complexifies the interpretation of “essentiality” of a gene. Additionally, some genes that are considered “essential” in a specific cell-line and experimental setting, might not appear as “essential” in biological or technical replicates (27). In this study, we thus considered diverse definitions of “essentiality” vs “non-essentiality”: we first used the “core essential genes” list from Bartha et al. (27). In their study, the authors discuss the discrepancies between conflicting definitions of “essentiality”: (i) genes whose inactivation can lead to embryonic lethality or severely reduced fitness, and (ii) genes intolerant to heterozygous loss-of-function variant as defined by the pLI. The first definition is centered around an individual, whereas the second one is defined at the populational level. By reconciling these two points of view, the authors provide a list of 1989 “core essential genes”. Second, we obtained lists of essential genes for specific cell-lines, established by CRISPR and/or RNAi experiments, from the Online GEne Essentiality database or OGEE versio 3.09 (https://v3.ogee.info/, (28)

Synthetic lethal (SL) gene pairs are defined as pairs of genes in which the simultaneous biallelic loss or inhibition of both genes (i.e. four alleles out of four) causes cell death, while the biallelic loss of either gene alone is compatible with cell viability (i.e. they are non-essential genes) (29). On the contrary, Synthetic Non-lethal (SNL) gene pairs are pairs of genes whose simultaneous loss or inhibition does not result in cell death. The same limitations as those in the definition of gene essentiality indicated above apply here, since they depend on the specific cell lines, experimental conditions, and replicates in which they were identified. The list of synthetic lethal pairs in human were obtained from the Synthetic Lethal Database (*synlethdb;* https://www.synlethdb.com/, version v3) was downloaded on 2024 February the 24^th^ (30,31). Only synthetic lethal pairs proven *in vitro* in specific cell-lines, involving CRISPR or RNAi experiments, were retained, whereas those predicted with bioinformatic tools or identified through text mining were filtered out. The synthetic lethal pairs reported in *synlethdb* come from different experiments, and as such, even though the same cell-lines are used in different studies, some other experimental conditions might not be identical, hindering the comparisons of the results. For the purpose of this work, we considered SL lists of specific cell lines. For the later, we retained the three cell lines with the highest number of SL pairs reported, i.e. K562 (a myelogenous leukemia cell line, https://www.cellosaurus.org/CVCL_0004), A375 (a melanoma cell-line, https://www.cellosaurus.org/CVCL_0132) and A549 (adenocarcinomic human alveolar basal epithelial cells, http://web.expasy.org/cellosaurus/CVCL_0023). In the previous definitions of SL pairs from specific cell lines, we filtered out the pairs of genes that were composed of at least one gene considered as “essential” according to strain-specific essential lists provided by OGEE (Online GEne Essentiality, https://v3.ogee.info/#/home) (28).

The list of all genes that are currently known to be involved in Monogenic Mendelian Diseases (MMDG) was obtained from OMIM (https://www.omim.org/), as described in Caron and Rausell, 2025 (32), where they used similar filtering steps as detailed in the paper from Chong and colleagues (33): OMIM data files were downloaded from https://www.omim.org/downloads/ on the 12/11/2024. Genes were mapped to their corresponding phenotypes, and the phenotypic descriptions were used to assess the corresponding genetic architecture of the phenotype: i) phenotypic description containing the keyword “somatic” were flagged as somatic, those containing the keywords “risk”, “quantitative trait locus”, “QTL”, “{”, “[” or “susceptibility to” were flagged as “complex”. We labelled as Mendelian disease genes those with a Clinvar supporting evidence level of 3 (i.e. disease molecular basis is known) and not flagged as somatic. These genes were further split in between monogenic mendelian disease genes (MMDGs, n= 4564) and complex mendelian disease genes (CMDGs, n=611) based on their “complex” flag. Among those, n=283 genes were associated with both monogenic and complex mendelian diseases. For the purpose of this work, we thus restricted the set of CMDGs to the 328 genes that are currently tagged as complex mendelian disease genes but not MMDGs.

The list of pairs of genes involved in human digenic disease was obtained from the benchmark introduced by Ogloblinsky et al. (12), based on the OLIDA v2.0 database (OLIgogenic DATAbase, https://olida.ibsquare.be/, 10.5281/zenodo.10732038) (6,7).

### Yeast knowledge-based biological network

#### Nodes

Analogous to the previous section, a yeast knowledge-based biological network was built in the form of a multiplex heterogeneous graph (**Supplementary Table 1**). This network integrates four distinct types of nodes and 14 edge types representing diverse biological relations. The four node types in the network represent protein-coding genes, and three categories of Gene Ontology terms (14,15).The first node type, hereafter referred as GENE, represents nuclear protein-coding genes for the reference yeast strain S288C (and/or the proteins they encode), and were obtained from the Saccharomyces Genome Database (SGD, https://www.yeastgenome.org/, accessed in June 2024, (34–37)). Genes located on the mitochondrial chromosome were excluded. The remaining three node types represent terms from the Gene Ontology (GO): Biological Process (BP), Cellular Component (CC), and Molecular Function (MF). These ontology terms and their relations were extracted from https://www.alliancegenome.org/bluegenes/alliancemine/results/ALL_Yeast_Genes, for the yeast strain S288C.

#### Edges

The network’s edges define relationships between the previous node types and consist of fourteenth distinct types (**Supplementary Table 1**). Of these, the Protein-Protein Interaction (PPI) and Paralogous edges are undirected; thus, each of these is represented by two directed edges in opposite directions, ensuring symmetry. As in the case of human, all other edge types are directed and were augmented with reverse edges to enable bidirectional message-passing operations during graph-based computations. Protein-protein interaction (PPI) edges connect pairs of GENE nodes and were sourced from version 12.0 of the STRING database (https://string-db.org/, released on July 26th 2023). Only high-confidence physical interactions between yeast proteins were retained (STRING combined score higher than 0.7). Paralogous edges also connect GENE nodes and were extracted from Ensembl release 112, released on May 13th, 2024, and downloaded on June 6th, 2024.). Gene Ontology relationships were extracted from https://www.alliancegenome.org/bluegenes/alliancemine/results/ALL_Yeast_Genes, for the yeast strain S288C, enabling the construction of three edge types: GENE-to-BP, GENE-to-CC, and GENE-to-MF. Each of these was paired with a corresponding reverse edge as previously indicated. In addition, GO ontology-internal relationships were represented by edges connecting BP terms, edges connecting CC terms, and edges connecting MF terms. Each of these was supplemented by a reverse edge. These hierarchical ontology relations were parsed from the go-basic.obo file provided by Gene Ontology. As in the case of human, the version used was format-version 1.2, released on November 3rd, 2022, and accessible at https://release.geneontology.org/2022-11-03/ontology/.

#### Node features

To characterize the 6579 protein-coding genes used as nodes in the yeast knowledge graph, we compiled a total of 17^th^ gene-level constraint and gene features. These metrics were integrated as numerical features for each GENE node, with harmonization of gene identifiers to ensure consistency across data sources. First, for each gene, we extracted the gene-level dN/dS ratio from Peter et al. (38). Second, gene-associated phenotypes observed in the S288C strain following deletion of individual genes, as reported in the Saccharomyces Genome Database (SGD, https://www.yeastgenome.org/, accessed in June 2024, (34–37)), were used as features for gene nodes in the knowledge graph. Among the 177 unique phenotypes catalogued, we selected only the sixteen phenotypes that were each associated with more than 1,000 protein-coding genes (excluding the phenotypes “inviable” and “viable”, which were used as node labels, see below): ‘chemical_compound_accumulation’, ‘chronological_lifespan’, ‘competitive_fitness’, ‘desiccation_resistance’, ‘haploinsufficient’, ‘heat_sensitivity’, ‘metal_resistance’, ‘oxidative_stress_resistance’, ‘replicative_lifespan’, ‘resistance_to_chemicals’, ‘respiratory_growth’, ‘stress_resistance’, ‘toxin_resistance’, ‘utilization_of_nitrogen_source’, ‘vacuolar_morphology’, ‘vegetative_growth’. Such phenotypes were one-hot encoded as boolean values, i.e. “1” (when a gene is associated to a given phenotype) and “0” otherwise.

#### Gene labels and pair-of-gene labels: gene essentiality and synthetic lethality

As in the case of human, we obtained several datasets allowing to assign GENE node labels and pair-of-GENE-nodes labels to the yeast knowledge graph, which will constitute the target labels we will aim to predict along the different GNN predictive tasks in this work, in the case of yeast, i.e.: gene essentiality and synthetic lethality. As for human, a gene was considered “essential” in yeast if its complete inactivation leads to cell death, whereas a gene is considered “non-essential” if its complete inactivation is tolerated. For this purpose, the phenotypes “inviable” and “viable” observed in the reference yeast S288C strain following deletion of individual genes, were obtained from the Saccharomyces Genome Database (SGD, https://www.yeastgenome.org/, accessed in June 2024, (34–37)).

The lists of synthetic lethal and synthetic non-lethal pairs in yeast were obtained from Costanzo et al. (39), where ∼23 million double mutants from Saccharomyces Cerevisiae were evaluated. Authors used synthetic genetic array (SGA) analysis, allowing the systematic construction of double mutants by crossing gene deletion mutant haploid yeasts in order to generate double-heterozygous diploid yeasts that generated 4 different types of cells by meiotic progeny (40). The haploid cells carrying both mutations can then be selected through a series of replica-pinning procedures and their fitness can then be scored. Genetic interactions involving “non-essential” genes were tested using gene knockouts, while those involving “essential” genes used thermosensitive mutants. For each gene pair, the authors measured the fitness of the single mutants and of the double mutant. The expected double mutant fitness (DMF) was calculated as the product of the two single mutant fitness values, assuming no interaction. The interaction score was defined as the difference between the observed and expected DMF. In Constanzo et al., a pair was defined as SL if the observed fitness was lower than expected, regardless of its absolute value. As a result, some SL pairs had higher observed fitness than pairs labeled SNL, due to smaller deviations from expectation. While this approach may be appropriate for detecting genetic interactions, we adapted the definition for the purpose of our study. Thus, to enable a more consistent generalization to digenism of the concept of gene essentiality, gene pairs with observed DMF scores were considered synthetic lethal in our study at varying thresholds (<0.5, <0.4, <0.3, progressively corresponding from a lesser to a more stringent definition), independently of their deviation of the expected DMF values.

### Betweenness centrality computation in the protein-protein-interaction networks

For both humans and yeast, betweenness centrality was computed using the NetworkX Python package (version 3.2.1). Betweenness centrality coefficients were calculated on a homogeneous graph composed exclusively of protein–protein interaction (PPI) edges. The PyTorch Geometric (PyG) homogeneous graphs were first converted into NetworkX Graph objects using the torch_geometric.utils.to_networkx function (https://pytorch-geometric.readthedocs.io/en/latest/modules/utils.html?highlight=to_networkx#torch_geometric.utils.to_netw <u>orkx</u>). Betweenness centrality was then computed using the networkx.betweenness_centrality() function (https://networkx.org/documentation/stable/reference/algorithms/generated/networkx.algorithms.centrality.betweenness_centrality.html) with default parameters.

### Graph Neural Networks

#### GNN architecture

Graph Convolutional Neural Networks are deep learning architectures able to handle graph-structured data to learn node representations, called node embeddings, leveraging both nodes features as well as the graph topology (41). A GNN model is composed of several Convolutional layers that will each compute such new node embeddings for each node by aggregating information from their neighbors. The design of convolutional layers for graph-structed data is adapted so that they are invariant to node ordering. To that aim, most GNNs use the so-called “message passing” framework, and are thus described as MPNN (Message Passing Neural Network). In this setting, each node sends a “message” (defined as a function on its features) to all its neighbors. Each node receives as well messages from all of its neighbors, and aggregates them using an aggregation function that is invariant to the node ordering, such as “mean”, “max” or “sum” functions. Diverse Graph Convolutional Layers can be used, depending on how they define the “message” and the “aggregation” functions. Several Graph Convolutional Layers can then be used sequentially to compute new node embeddings. A non-linear activation function, i.e. ‘relu’ or ‘tanh’, is used between each graph convolutional layer in order to learn more complex representations. After the final nodes embeddings have been computed, a classical neural network architecture, with a linear/fully connected layer is used to perform downstream predictive tasks. Loss function used throughout this work, including both node and edge prediction tasks, was binary cross entropy and optimization was performed using Adam algorithm with a learning rate of 0.01 and a weight decay of 0.0005 (42,43).

The notion of “receptive field” is used to represent the set of all the nodes from which a given node u will aggregate information in order to obtain its final embedding. Concatenating different graph convolutional layers allows to gather information from nodes that are further “hops-away” in the graph. Thus, the initial embedding of a node is simply its nodes features and is written 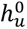. A graph convolutional layer is then used in a message-passing framework to compute new nodes embeddings for each node by aggregating information from their direct neighbours. The layer-1 embedding of a given node is written 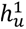 and depends on the level-0 embedding (i.e. initial node features) of its neighbours. Concatenating 2 graph convolutional layers allows to compute level-2 embeddings for each node. For a given node u, its level-2 embedding 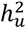 depends on the layer-1 embeddings of its neighbors, that, in turn, depends on the level-0 embeddings of their neighbors, so information from the neighbors of the neighbors of node u are needed to compute its level-2 embeddings. More generally, the final embedding of a node in a GNN composed of n-layers will depend on all the nodes that are separated by fewer than (n+1) connections. The set of all these nodes that are closer than “n-hops away” from node u is called the receptive field of node u.

The first graph convolutional layer developed was the GCN (Graph Convolutional Network) convolution, which uses a weighted sum of the neighbors’ features plus the node’s own features to compute embeddings (213). The weight matrices are learnt during training. In our work, we used two more complex Graph Convolutional Layers: GATConv and SAGEConv (44–46). GATConv (Graph Attention) uses attention coefficients specific to each edge to perform more expressive aggregations (44,45), i.e. allowing that the different nodes composing the neighborhood of a specific node have different relative importance. Thus, more important nodes should be given a higher attention coefficient in order to increase the performance of the model, and the number of “attention heads” can be specified in order to increase model complexity. In SAGEConv, different functions such as ‘mean’, ‘add’, etc. can be used to aggregate the neighbors’ features, and a normalization of the features and other parameters can be applied (46).

GNNs can be adapted to handle heterogeneous graph-structed data, i.e., when several node types and/or node edges are present. Thus, in our work, heterogeneous GNNs were built by defining specific graph convolutional layers for each edge, which will compute edge-specific embeddings that are then aggregated using a rotation invariant aggregation function such as ‘mean’, ‘max’ or ‘sum’. Throughout this work we used the aggregator ‘sum’ in order to group the node embeddings that were generated by different relations.

### GNN computational framework

This work was done using the programming language Python version 3.10.14. The main package used was PyTorch Geometric (47,48) which is a wrapper around Pytroch, developed in order to handle graph-structured data. A conda environment was created using the conda package (version 23.3.1) and the following packages were installed at the creation of the environment in order to allow a proper Pytorch installation: cuda-nvcc version 11.8.89, cudatoolkit version11.8.0, cudnn version 8.9.2.26. After installation of these 3 packages, installed, the following packages were installed: Numpy version 1.26.3, Pandas version 2.2.1, Scikit-learn version 1.4.1, Torch version 2.1.2 installed with CUDA 11.8: 2.1.2+ptcu118, Torch-cluster version 1.6.3, Torch-scatter version 2.1.2, Torch-sparse version 0.6.18, Torch-spline-conv version 1.2.2, Torchaudio version 2.1.2, Torchvision 0.16.3, Torch-geometric version 2.5.2, pyg-lib version 0.3.1, Goatools version 1.3.1, Pybiomart version 0.2.0, Intermine version 1.13.0, Jupyter version 1.1.1. Scripts were run on a Linux server (CentOS Linux 7, Kernel: Linux 3.10.0-1160.119.1.el7.x86_64) with 72 cores, 630 Go of RAM and 2 GPU RTX 2080 Ti

Heterogeneous GNNs were computed using the HeteroConv wrapper from PyG (https://pytorch-geometric.readthedocs.io/en/latest/generated/torch_geometric.nn.conv.HeteroConv.html#torch_geometric.nn.conv.HeteroConv) which is a dictionary-like-structured object that is used to define GNN models applicable to heterogeneous graphs by defining edge-specific graph convolutional layers. A 2-layers GNN applied to homogeneous graphs is composed of only 2 graph convolutional layers, whereas a 2-layers GNN applied to heterogeneous graph is composed of 2 HeteroConv layers, each of them being composed of as many graph convolutional layers as they are types of edges in the graph. The GATv2Conv or the SAGEConv graph convolutional layers were used for all edge-types in our models. The GATv2Conv was used with the default parameters except for the ‘add_self_loops’ argument which was set to ‘False’ for GENE-to-GENE connecting edges, except for edge prediction tasks, and the ‘heads’ parameter was set to either 1 or 2 (369,370). The SAGEConv was used with parameters ‘aggr’ set to ‘sum’, the ‘normalize’ parameter set to True and the ‘root_weight’ parameter set to False for GENE-to-GENE, GENE-to-not-GENE and not-GENE-to-GENE connecting edges in the first HeteroConv layer, except for edge prediction tasks (371).

### GNN architectural adaptations to classify pairs-of-nodes as new nodes: the PAirs of Genes As geNes (PAGAN) approach

In this work we propose to infer pair-of-gene labels by training a given GNN model to predict individual node labels and apply it to classify an independent set of pairs of nodes, where pairs of nodes are treated as if they were additional new individual nodes in the knowledge graph network. We named this approach as PAGAN: “**PA**irs of **G**enes **A**s ge**N**es” approach. The following GNN architectural adaptations were implemented in PAGAN:

First, modelling pairs as new nodes required to create new nodes as well as new edges in the Knowledge Graph. The node-type of such new nodes was considered be the same as the node-type of the nodes composing the pairs, and these new nodes were connected to all the neighbors of the two nodes composing the pair. Let’s write *N_u_* the neighborhood of node *u*, which is composed of all the nodes connected to node *u* where *u* is the destination-node of the corresponding edges. Similarly, we can note *N_v_* the neighborhood of node v. We then defined *N_u+v_* as the neighborhood of the node *u+v*, which is composed of all the nodes in the neighborhood of node *u* and all the nodes in the neighborhood of node *v*. The rationale of this approach is that, if gene *u* is associated with a biological entity X (i.e. node) by virtue of a given type of relation (i.e. edge), then the pair of genes *(u+v)* can be assumed to be associated with entity X by virtue of the same relation type. We assumed this for all relations (edges) in the knowledge graph described above except for the ‘expressed in’ relation associating ‘GENE’ nodes with ‘ANATOMY’ nodes. In the later relation, the new node representing the pair (*u+v*) was connected to a node of type ‘ANATOMY’ only if such node was connected to both gene *u* and gene *v*. Thus, if gene *u* is expressed in tissues T1 and T2, and that gene *v* is expressed in tissues T1 and T3, then the “gene” (*u+v*) was only considered to be expressed in tissue T1.

Second, in the PAGAN approach we imposed that a node (*u+v)* representing a pair was not itself connected to any of the original genes *u* and *<u>v</u>* (i.e. nodes), even if *u* and *v* were themselves connected by a given edge in the knowledge graph. In such a way, we ensure that the prediction of a node representing a pair is not dependent on the features of the genes composing it.

Third, we imposed as well that all the edges connecting the pre-existing nodes in the knowledge graph to the new nodes representing pairs, are considered as undirected edges, in a way that the nodes representing pairs are always “destination nodes”, and not “source nodes”. This was imposed for all relations (edges) including those that are reciprocal in the original knowledge graph, such as protein-protein-interactions. This constraint ensures that, in the message-passing framework, messages will only flow from the pre-existing nodes to the new nodes representing pairs, and not in the opposite direction. Thus, while the new nodes representing pairs will aggregate information from the original nodes, the original nodes will not use messages passed thought nodes representing pairs when computing their embeddings.

Finally, we modified the default behavior of GNNs regarding the node embedding update in the graph at each graph convolution, where information from their neighbors is aggregated at each layer in the message-passing framework. By default, the embedding at the layer *(n+1)* for node *u* is computed using the features/embeddings at the layer *n* for node *u* and all of its neighbors. However, in PAGAN, GNN models were implemented in a way that the embeddings at layer *(n+1)* for node *u* were computed using only the features/embeddings at the layer *n* of all of its neighbors, but not those of node *u* itself. This is done both during training as well as at inference time. This allowed PAGAN to make predictions on nodes representing pairs of genes, for which there were no analogous node features to nodes representing individual genes. In the context of the Pytorch Geometric library, this was achived by setting the parameters “add_self_loops” (for the GATv2Conv) or “root_weight” (for the SAGEConv) to False (https://github.com/pyg-team/pytorch_geometric/discussions/7408). Yet, when using at least two graph convolutional layers with undirected edges, a node’s initial features can propagate back to itself within the message-passing framework. This happens because each node belongs to its own 2-hop neighborhood. Consequently, the embedding of a node u at layer n+2 depends on the embeddings of its neighbors at layer n+1, which in turn depend, at least partially, on the embedding of node u at layer n.

To prevent a node’s initial features from influencing its own embedding after two convolutional layers, we slightly modified the message-passing process. This was achieved using the NeighborLoader class in the PyTorch Geometric (PyG) library (https://pytorch-geometric.readthedocs.io/en/latest/modules/loader.html?highlight=neighborloader#torch_geometric.loader. NeighborLoader). In its default setting, this function divides the dataset into different batches, each of them defined by its set of seed nodes (or anchor nodes) whose number is defined by the ‘batch_size’ parameter. The NeighborLoader will then create a subgraph centered around the seed nodes of the batch by taking all the neighbors required to compute the nodes embeddings. This will include all the direct neighbors, i.e. all the nodes that are connected to the anchor nodes by a direct edge (1-hop away), as well as the indirect neighbors, i.e. those connected to the anchor nodes by a path passing through several nodes (2 or several-hops away). The sampled subgraph is dependent of the ‘batch_size’ and the architecture of the model, the higher the number of graph convolution layers used in the model, the larger the size of the subgraph sampled. In the case of heterogeneous graphs, it is possible to further specify different parameters such as the number of neighbors that need to be sampled per edge type and per layer. However, in PAGAN we set the ‘disjoint’ argument of the PyG NeighborLoader to True, so that each seed (or anchor) node in the Neighborloader generates its own disjoint subgraph, rather than a single global subgraph that includes all neighbors of all anchor nodes in the batch. Within each subgraph, we thus zero out the initial features of the central (anchor) node. This ensures that the node’s final embedding is not dependent on its own features, while not impacting the final embeddings of the other nodes in the batch. For example, with a batch size of 2, the NeighborLoader will create two separate subgraphs centered around nodes u and v, respectively, where u and v are connected. Node v will appear in u’s subgraph and vice versa. In the subgraph centered on u, node u’s features are zeroed out, while node v’s features are used normally. Conversely, in the subgraph centered on v, node v’s features are zeroed out, while node u’s features remain intact. This approach increases memory usage and computation time, since nodes can appear in multiple subgraphs, but it allowed us to zero out the anchor nodes’ features in each batch (https://github.com/pyg-team/pytorch_geometric/discussions/7408). Importantly, while a node’s features are not used for its own embedding, they still participate in the message-passing process that determines the embeddings of its neighbours.

### GNN training, validation and testing across node prediction and edge prediction tasks

Throughout this work, inductive GNN learning was performed, i.e. where models are trained on a subset of nodes/edges and can be applied to different nodes/edges that were not labelled/present at training time. Two different prediction tasks were performed: (1) node classification tasks, where the goal is to predict the class of a given nodes; here the model is trained on a subset of the nodes and tested on the others. And (2) edge prediction task, where the goal is to predict node connections in the graph. It is worth noting that, in this situation, the final prediction for each edge was computed using the neighborhood’s information as well as the nodes’ own features connected by the edge. Splitting graph-structured data into train, validation and test sets depends on the classification task. For node-classification, as previously stated, we used the NeighborLoader object from the Pytorch Geometric package. In the case of edge prediction tasks, edges need to be divided into a train, validation and test set, rather than individual nodes. However, these edges can also be used by the graph convolutional layers in the message passing framework, so the split must be carefully designed as follows: In a homogeneous graph (with only one node-type and only one edge-type), some edges will be removed from the graph and the model will be trained to predict them whereas the remaining edges will be used as message-passing edges. The removed edges will be used as “supervision edges” and will be associated with the target 1 whereas false edges will be randomly sampled from the graph and will be associated with the target 0. The RandomLinkSplit function from the pyg library can split the initial graph into a train, validation and test sets (https://pytorch-geometric.readthedocs.io/en/latest/generated/torch_geometric.transforms.RandomLinkSplit.html?highlight=randomlinksplit#torch_geometric.transforms.RandomLinkSplit). These sets are concentric, with the inner-most one (i.e. the train set), containing the least number of edges and the outermost-one, the test set, containing all the edges. All the message-passing edges in an outer set are composed of both the message-passing edges and the “supervision edges” that were in the inner set. The “supervision edges” of this outer set were left-out and not seen in any inner set. In order to avoid data leakage between the train and test sets, we set that edges to be predicted are undirected, which will functionally divide by 2 the number of edges to be used in supervision. In order to perform batch learning, we used the LinkNeighborLoader function (https://pytorch-geometric.readthedocs.io/en/latest/modules/loader.html?highlight=linkneighborloader#torch_geometric.loader.LinkNeighborLoader).

Throughout the work, models were trained for a maximum of 500 epochs, and were prematurely stopped when the train loss didn’t decrease significantly for 10 consecutive epochs (i.e. train loss decrease tailored to either 0.01 or 0.005 depending on the specific model’s computing time).

### PAGAN node-classification training and testing scenarios: genes-to-genes (g2g), pairs-to-pairs (p2p) and genes-to-pairs (g2p)

To evaluate the PAGAN approach, three different experimental settings were considered involving GNN node classification tasks:

**(i) Training and testing on individual genes**, hereafter referred as the genes-to-genes (g2g) setting. In this scenario, node classification tasks involved first gene essentiality and non-essentiality prediction, both for yeast and human (see sections “Node labels and pair-of-nodes labels”, above, for both human and yeast). In the case of human, the “core essential genes” list from Bartha et al. (27) was used. For both organisms, we randomly split the target nodes into a train and test sets with a 70%/30% ratio using 30 different random seeds. In addition, in the case of human, an additional node classification task was evaluated consisting in the discrimination of Monogenic Mendelian Disease Genes (MMDG) from non-disease genes (see section “Node labels and pair-of-nodes labels” of the human knowledge graph described above). The individual genes were split into 10 genomic partitions, equally stratified across labels, and models were trained on 9 partitions in order to be tested on the last one that was left-out.
**(ii) Training and testing on pairs of genes**, represented as if they were individual genes, hereafter referred as the pairs-to-pairs (p2p) approach. In this scenario, pairs of genes were introduced in the knowledge graph as new nodes representing the pair, and subsequently treated as nodes by the GNNs (see **section “GNN architectural adaptations to classify pairs-of-nodes as new nodes”**). Node classification tasks involved synthetic lethality (SL) and synthetic non-lethality (SNL) prediction, both for human and yeast (see sections “Node labels and pair-of-nodes labels”, above, for both human and yeast). In the case of human, only SL pairs from the K562 cell line were considered in the pairs-to-pairs approach, as those from the A375 and A549 cell lines were not numerous enough for train/test splitting in this setting. To avoid any overlap between the training and test sets, either in terms of genes or of pairs of genes, the following steps where carried out. Both for human and for yeast, the individual genes composing the SL pairs were split into a training gene set and a testing gene set with a 80%/20% ratio. SL pairs composed of 2 genes belonging to the training gene set were used as SL within the training set, while SL pairs composed of 2 genes belonging to the testing gene set were used in the testing set. SL pairs composed of one gene belonging to the training gene set and one gene belonging to the testing gene set, were filtered out. In the case of human, SNL pairs were generated by randomly sampling two different subsets of non-essential genes (as defined above), by using two different strategies, referred as (i) matched and (ii) unmatched. In the matched SNL set, for each randomly generated SNL pair, one gene was sampled from the set of individual non-essential genes composing the SL pairs (as defined above), whereas the other one was sampled from the set of individual non-essential genes that were not found in SL pairs. On the contrary, in the unmatched SNL, both genes were sampled from the set of individual non-essential genes that were not found in SL pairs. This was done in order to evaluate to which extent the performance of the GNN model depends on the type of individual genes composing the SNL pairs. In the case of yeast, SNL pairs were sampled from the datasets described above, retaining pairs with DMF between 0.99 and 1.01, and an interaction score (ε) below 0.1. Out of the 6579 yeasts genes, there were a total of 1311 essential and 5268 non-essential genes. Among the 5268 non-essential genes, only 4262 were assessed in Constanzo et al, and classified as either SL (as defined by the DMF criterion above) or SNL pairs. Of note, all SNL pairs were composed of genes that were also participating in SL pairs, except for the gene *PTK1* (systematic name: YKL198C) which was only found in SNL pairs. For yeast, all 4262 individual non-essential genes were considered when retaining SL and SNL pairs; such individual genes were split into 80%/20% sets to define training and testing gene sets, respectively. In this setting, a remaining set of 1006 individual non-essential genes were not used in any SL or SNL pairs.
**(iii) Training on individual genes and predicting on pairs of genes**, representing such pairs as if they were individual genes, hereafter referred as the genes-to-pairs approach (g2p). In this setting we evaluated whether different GNN approaches each trained to classify genes according to a give biological category (i.e. gene essentiality/non-essentiality, or Monogenic Mendelian disease gene (MMDG)/non-MMDG) could generalize to classify their digenic counterparts, i.e. SL/SNL pairs or disease/non-disease gene pairs, respectively. We did so both on human and yeast. In this setting, we re-trained the analogous GNN models for individual gene classification described in the genes-to-genes setting (see above). However, in this case, instead of splitting the labelled nodes into train/test sets, the entire set of labelled nodes may be used for training, as the models are now evaluated on gene pairs. The retrained GNN models were then evaluated on the corresponding gene pairs described in the pairs-to-pairs setting (see above). Similarly, rather than splitting the labelled pairs into train/test sets, the entire set of labelled pairs (represented as new nodes) was used for testing, as the models are now trained on individual genes. In the case of yeast, two training settings were evaluated: in the first, PAGAN models were trained to classify the 1311 essential and 5268 non-essential genes in the knowledge-graph. In a second setting, the non-essential genes participating in SL or SNL pairs were excluded from the training set, i.e. minimizing the risk of data leakage between train and test sets under the genes-to-pairs setting. This led to a training-set in the genes-to-pairs experiments composed of set of 1311 essential genes and 1006 non-essential genes that were not part of either SL or SNL pairs (see above.

In the case of human, for the purpose of the analyses in the genes-to-pairs approach, the GNN model to classify genes according to gene essentiality/non-essentiality described above, was trained on lists of essential genes for specific cell-lines, established by CRISPR, from the Online GEne Essentiality database or OGEE version 3.09 (https://v3.ogee.info/. Cell lines considered were again K562, A375 and A549, i.e. the ones with the highest number of SL pairs reported in *synlethdb*. Two settings for the PAGAN training were considered: one considering all essential and non-essential genes for a given cell line, and another one excluding from the training set the non-essential genes participating in SL or SNL pairs. For testing purposes, the corresponding cell-line specific lists of SL pairs were considered, as described above. We generated SNL pairs by random sampling as previously defined in the pairs-to-pairs approach (see above). All the SL pairs as well as the ‘matched’ SNL pairs and ‘unmatched’ SNL pairs built in the pairs-to-pairs setting (see above) were used in the corresponding test sets, leading to a 2:1 ratio between SNL and SL pairs.

In humans, an additional variation of the genes-to-pairs (g2p) setting was tested on pathogenic and non-pathogenic gene pairs. Here we trained the PAGAN approach on the task of discriminating n=4,541 Monogenic Mendelian Disease Genes (MMDGs) from n=14,362 non-disease genes, as described above, and evaluated these models on its ability to discriminate a curated set of n=255 unique human disease gene pairs from n=204,583 unique non-disease gene pairs derived from the Ogloblinsky benchmark dataset (12), which is based on the OLIDA database (6,7). The Ogloblinsky et al. benchmark dataset consisted of pairs of variants involving two genes associated with genetic diseases, whereas the negative dataset comprises variant pairs involving two genes not implicated (as a pair of variants) in genetic disorders.

### Hyperparameter Selection in the PAGAN models

The following hyperparameters were initially explored in the previously described PAGAN genes-to-genes setting for gene essentiality and non-essentiality prediction, on both yeast and human: the convolution type (SAGEConv or GATv2Conv), the number of convolutional layers (1, 2, or 3), the activation function (ReLU or tanh), the embedding dimension (16, 32 or 64), and the batch size (16, 32, 64, or 128). The Area Under the Receiver Operating Characteristic (AUROC) and the Area Under the Precision–Recall Curve (AUPR) were first evaluated through 10-fold cross validation on one of the random train splits, representing 70% of the target nodes (**Supplementary Figure 1**). From such first exploratory evaluation, it was observed that models using GATv2Conv consistently underperformed compared to SAGEConv. Consequently, GATv2Conv was excluded from further tests. Similarly, models with an embedding dimension of 128 showed no performance improvement, but required substantially higher computational time and were therefore discarded. Increasing the number of graph convolutional layers from two to three did not yield significant performance gains either but markedly increased training time. Conversely, using only one layer accelerated training but degraded performance. Thus, two graph convolutional layers were selected as the optimal trade-off between accuracy and efficiency. Regarding batch size, models trained with a batch size of 128 consistently outperformed smaller batch sizes (16, 32, and 64). Based on these results, subsequent experiments were conducted using SAGEConv as the convolution type, two graph convolutional layers, and a batch size of 128. The remaining hyperparameters retained for tuning were the activation function (ReLU or tanh) and the embedding dimension (16, 32, or 64). This configuration reduced the search space to six possible combinations. Subsequently, all models in the different PAGAN experimental settings were trained using these six hyperparameters combinations, and model performance was evaluated using AUROC and AUPR values.

In addition, in the previously described PAGAN genes-to-genes setting for human gene classification into Monogenic Mendelian Disease Genes (MMDG) from non-disease genes (see section “Node labels and pair-of-nodes labels” of the human knowledge graph described above). GNN models used two *HeteroConv* layers, using SAGEConv for each edge-type and a batch size of 128. We trained six models on each train set correspondingto six hyperparameter combinations involving different embedding size (16, 32 or 64) and non-linear activation functions (‘relu’ or ‘tanh’).

### Comparison with alternative state-of-the-art methods for gene classification

For gene (i.e. node) classification tasks, our approaches were compared with alternative supervised-learning methods using either gene features, network-based characteristics, or both: Logistic Regression models were implemented using the ‘LogisticRegression’ class from Scikit-learn (https://scikit-learn.org/stable/modules/generated/sklearn.linear_model.LogisticRegression.html). Hyperparameter optimization through cross-validation was performed on the following parameters: tolerance for stopping criteria (“tol”) values of 0.0001, 0.001, 0.01, and 0.1; inversed of regularization strength (“C”) values of 1.0, 2.0, and 3.0; and algorithm to use in the optimization problem (“solvers”) set to either ‘’lbfgs’’ or ‘’newton-cholesky’’. An Artificial Neural Network (ANN) was also implemented, consisting of two fully connected hidden layers with an embedding size of 16 and using the ReLU activation function. Additionally, a Metapath2Vec model (https://pytorch-geometric.readthedocs.io/en/latest/generated/torch_geometric.nn.models.MetaPath2Vec.html#torch_geometric.nn.models.MetaPath2Vec) was employed as a random-walk (RW) based approach applicable to heterogeneous graphs. The following hyperparameters were used: embedding dimension of 64, walk length of 100, context size of 10, twenty walks per node, and five negative samples per positive context. For graph-based learning, a classical Graph Neural Network (GNN) with two graph convolutional layers using ‘SAGEConv’ was trained, with an embedding size of 16 and the ReLU activation function. In parallel, our modified GNN architecture was implemented with the same configuration—two convolutional layers using ‘SAGEConv’, embedding size of 16, and ReLU activation, but with adjustments described previously to prevent nodes from using their own features when updating embeddings.

In the Logistic Regression and ANN models, predictions for each gene relied exclusively on their individual gene features. In contrast, predictions from the Metapath2Vec model were derived solely from random walks through the heterogeneous network and did not incorporate node features. The classical GNN utilized both node features and those of neighbouring nodes to compute final embeddings and predictions.

### Comparison with alternative state-of-the-art methods for gene pairs classification: synthetic lethality and digenism prediction

For synthetic lethality (SL) prediction in human, our approaches were compared with SLGNN, a supervised graph neural network (GNN)–based method for edge prediction that utilizes the knowledge graph provided by SynLethDB. SLGNN was previously reported to outperform alternative state-of-the-art methods for SL prediction and was considered a baseline in our study (49). The SLGNN dataset consists of 36,042 SL pairs extracted from SynLethDB and 36,402 randomly generated non-synthetic-lethal (SNL) pairs originally used by the KG4SL method (50). Model training and evaluation were conducted using five-fold cross-validation. During this process, certain gene pairs were duplicated across some of the training, validation, and test splits to increase dataset size. As a result, overlapping pairs were present across these sets, which could introduce data leakage and inflate performance estimates. Additionally, several gene identifiers in the SLGNN dataset corresponded to non-human genes (e.g., Mus musculus), and some pairs included non–protein-coding genes, such as pseudogenes or long non-coding RNAs. Moreover, some genes included in the SLGNN SL pairs were annotated as essential genes in other datasets used in this study. These issues made the original SLGNN model less comparable to our approach. Thus, for the purpose of comparison in our work and to ensure consistency across methods, the SLGNN model was retrained on a reduced dataset containing only gene pairs in which both genes were present in both our knowledge graph and the SLGNN knowledge graph. Retraining of SLGNN was performed using the publicly available implementation (https://github.com/zy972014452/SLGNN), using its original DGL-based conda environment, without any modification to the provided scripts. Only the input files (train.txt, val.txt, and test.txt) were replaced with the corresponding gene identifiers from our datasets. While the SLGNN knowledge graph is sparser than ours, it contained more edge types. SLGNN does not incorporate node features directly; instead, it generates node embeddings from the knowledge graph, which are subsequently used as input features for a second graph. This design enables efficient computation; however, because the embedding learning and downstream training graphs are linked, the backpropagation process renders the embedding learning supervised, meaning that the learned embeddings may not be entirely unbiased representations of the underlying graph structure.

For disease gene pair prediction, our approaches were compared with alternative state-of-the-art methods following the benchmark implemented in Ogloblinsky et al. (12), which was based on the OLIDA database (6,7). Methods evaluated included Varcopp2.0 (10), DiGePred (8), DIEP (9) and ARBOCK (11). In the benchmark, all tools were assessed on a common test set without retraining their original implementations (see above). Methods preceding Varcopp2.0 (10), such DiGePred (8) and DIEP (9), had been trained on smaller subsets of digenic pairs, while ARBOCK (11), developed subsequent to Varcopp2.0. Accurately determining the training set used by each of these tools was challenging, and could not be confirmed whether the Ogloblinsky et al. benchmark test set included only gene pairs whose individual genes were fully disjoint from those present in their training set, except in the case of Varcopp20.

## Results

### 1. The PAGAN approach

In this work we implemented a modified Graph Neural Network (GNNs) approach for the classification of pairs of genes by mining biological relations represented in the form of a knowledge graph (**Figure 1**; **Methods**). Graph Neural Networks (GNNs) are a machine-learning approach suitable to classify entities represented as nodes in a graph by iteratively aggregating information from their neighbors, thereby combining node features with the structure of the underlying network. Here, genes are embedded in a heterogeneous biological knowledge graph together with functional annotations (e.g. biological processes, molecular function and tissue expression, among others), and a GNN is first trained to predict labels for individual genes based solely on their network context. We extended this principle to gene pairs by representing each pair of genes as a new artificial node in the same graph. We thus called the approach as PAGAN approach (PAirs of Genes As geNes). In the PAGAN GNN implementation, a pair node inherits the combined neighborhoods of the two genes, meaning it is connected to all biological entities linked to either gene, while respecting relation-specific constraints. Importantly, the pair node is not connected to the two genes themselves, and information flow is constrained so that messages propagate only from existing biological nodes to the pair node. As a result, predictions for gene pairs rely exclusively on their shared and combined network context, and not on the intrinsic features of the individual genes. By training a GNN on single-gene labels and then applying it unchanged to these pair nodes, PAGAN enables the inference of pair-level properties, such as synthetic lethality or digenic disease, from patterns learned at the gene level. This strategy allows biological characteristics of gene pairs to be inferred without explicitly training on large sets of labeled gene pairs, providing a unified framework for generalizing genes properties to gene combinations.

**Figure 1.**
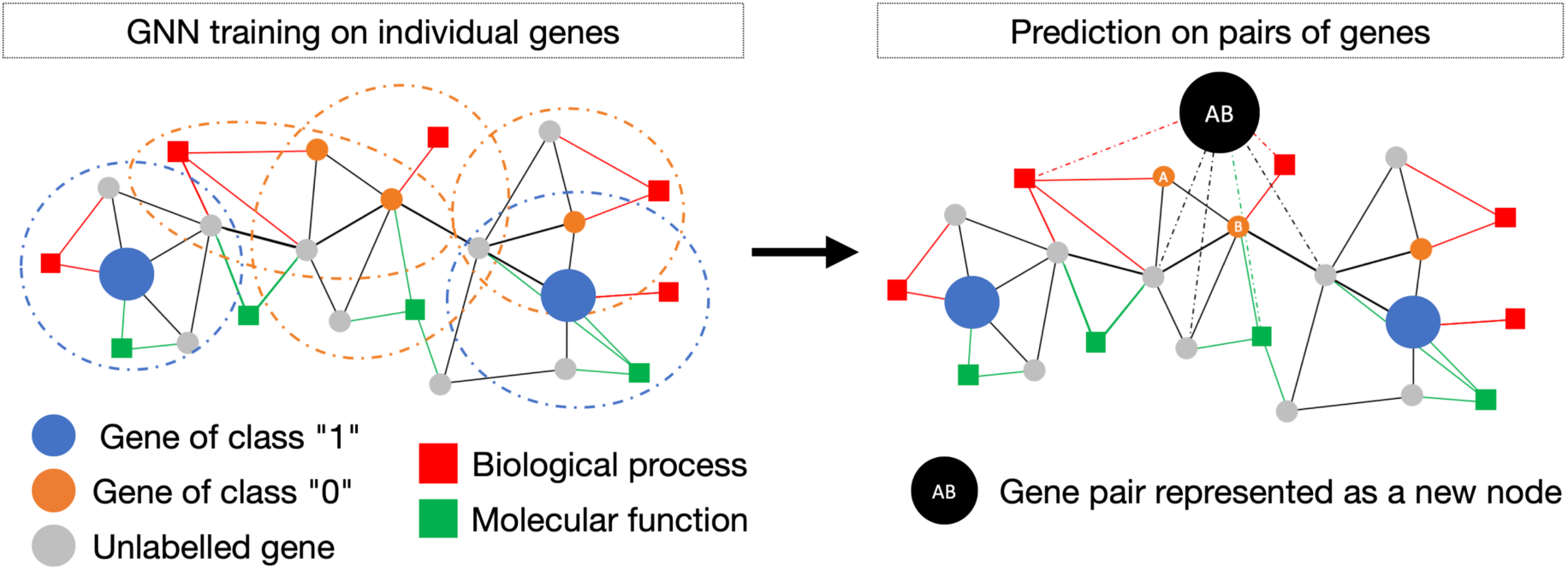
Schematic overview of the PAGAN approach (PAirs of Genes As geNes). **Left**: A Graph Neural Network (GNN) is trained to classify individual genes in a heterogeneous biological knowledge graph by aggregating information from their neighboring biological entities (e.g. biological functions, molecular processes, among others). **Right**: To enable prediction on gene pairs, each pair is represented as a new node in the graph, connected to the union of the neighbors of the two genes. Pair nodes are not connected to the original genes, and information flows only from existing nodes toward the pair node. The trained GNN is then applied unchanged to these pair nodes, allowing inference of pair-level properties from their combined network context.

### 2. Gene essentiality and synthetic lethality

We first evaluated the PAGAN approach by its capacity to learn biological network characteristics associated with essential genes and with synthetic lethal gene pairs. We did so both in yeast and human, independently. Yeast was a particularly suitable model organism for the purpose of our study, as both gene essentiality as well as the fitness effect of virtually all possible double gene mutants have been systematically studied. Two knowledge-based biological networks, for yeast and human respectively, were built in the form of multiplex heterogeneous graph. These networks integrate distinct types of nodes and edge representing diverse biological relations (**Methods**, **Supplementary Table 1**).

#### 2.1 Learning on individual genes, predicting on individual genes: the genes-to-genes setting

We first applied the PAGAN approach to the training and testing on individual gene essentiality, referred as the genes-to-genes (g2g) setting (**Methods**). In the yeast knowledge graph, a total of 1311 genes were classified as essential and 5268 as non-essential (**Methods**). We randomly split the yeast knowledge graph into a train and test sets with a 70%/30% ratio using 30 different random seeds. For each of the 30 random splits, each training set of 4604 genes was further divided into 10 folds for cross-validation, with nine folds (4143 genes) used for training and one fold (461 genes) reserved for validation. Across all cross-validation folds and random seeds, the models achieved an average validation AUROC of 85.35±1.19 % and an average AUPR of 67.92±2.44 %. On the left-out test sets, the average performance across all random seeds was AUROC = 84.89±1.36% and AUPR = 68.42±2.28% (**Table 1**). As a reference, the equivalent classification performed by using the gene’s betweenness centrality in the protein-protein interaction network of the knowledge-graph reached to an AUROC = 71.44% and AUPR = 37.11%.

**Table 1.** Comparative benchmark of the PAGAN genes-to-genes approach on the classification of gene essentiality. The table reports the average and standard deviation (std) of the AUROC and AUPR obtained across the left-out test sets obtained through alternative random-splits (Methods). The results are reported for the PAGAN genes-to-genes approach together with alternative baseline models trained on the same gene features and/or network edges, including: a Logistic Regression model, a classical Artificial Neural Network (ANN), the Metapath2Vec model, and a conventional Graph Neural Network (GNN). Further details are provided in the text and in Methods.

| Organism | Classifier | AUROC | AUPR |
| --- | --- | --- | --- |
| Yeast | PAGAN genes-to-genes setting | 84.89% (std: 1.36%) | 68.42% (std: 2.28) |
|  | Classical GNN | 93.45% (std: 0.60%) | 87.18% (std: 1.10%) |
|  | ANN | 69.29% (std: 1.20%) | 28.66% (std: 0.90%) |
|  | Logistic Regression | 70.99% (std: 0.96%) | 29.53% (std: 0.76%) |
|  | Metapath2Vec | 75.22% (std: 1.60%) | 44.94% (std: 2.26%) |
| Human | PAGAN genes-to-genes setting | 90.10% (std = 0.73%) | 63.20% (std = 2.28%). |
|  | Classical GNN | 91.73% (std: 0.63%) | 66.46% (std:1.81%) |
|  | ANN | 63.19% (std: 1.34%) | 17.60% (std:0.92%) |
|  | Logistic Regression | 64.33% (std: 1.20%) | 19.48% (std: 0.90%) |
|  | Metapath2Vec | 88.64% (std: 0.56%) | 59.28% (std: 1.83%) |

To further assess performance, our approach was compared against several baseline models trained on the same gene features and/or network edges (**Table 1**; **Methods**). A Logistic Regression model achieved on average an AUROC of 70.99%, while a classical Artificial Neural Network (ANN) composed of two hidden layers with 16 neurons each reached an average AUROC of 69.29%. The Metapath2Vec model, based solely on random-walk–derived embeddings, achieved an AUROC of 75.22%. A conventional Graph Neural Network (GNN) using node features during message passing outperformed all other models, reaching an AUROC of 93.28%. As previously indicated, in the modified GNN architecture of the PAGAN approach, node predictions rely exclusively on neighbouring features.

In the case of human, the knowledge graph contained a total of 1989 essential and 17341 non-essential genes (**Methods**). As in the case of yeast, we randomly split the yeast knowledge graph into a train and test sets with a 70%/30% ratio using 30 different random seeds. For each of the 30 random splits, each training set of 13530 genes was further divided into 10 folds for cross-validation, with nine folds (12176 genes) used for training and one fold (1354 genes) reserved for validation. Across all cross-validation folds and all random-seeds, the average performances of the models on the validation sets were: AUROC = 90.07±0.43 % and AUPR = 62.91±1.14 %. Across all random-seeds, the average performances of the models on the left-out test set were: AUROC = 90.10±0.73 %, and AUPR = 63.20±2.28 %. As a reference, the equivalent classification performed by using the gene’s betweenness centrality in the protein-protein interaction network of the knowledge-graph reached to an AUROC = 72.51% and AUPR = 21.37%. Analogous to yeast, our approach was compared against several baseline models trained on the same gene features and/or network edges (**Table 1**, **Methods**). Similar trends were generally observed in humans, although in this case the two versions of the GNN approaches achieved comparable performance, suggesting that the gene’s neighborhood provided strong predictive power, even when its individual features were disregarded.

Considered together, these results confirm that the GNN-based architectures can effectively exploit heterogeneous biological relationships to achieve more accurate and robust predictions than traditional machine learning methods or standard ANNs. In addition, the modified GNN version implemented in the PAGAN approach, disregarding the gene-target features, yielded slightly lower or comparable performances than the classical GNN, while having the advantage of being applicable to nodes lacking intrinsic features.

#### 2.2. Learning on pairs of genes, predicting on pairs of genes: the pairs-to-pairs setting

We then evaluated the PAGAN approach by its capacity to learn biological network characteristics associated to synthetic lethal gene pairs. We thus applied PAGAN to the training and testing on synthetic lethal pairs, referred as the pairs-to-pairs (p2p) setting (**Methods**). In this scenario, pairs of genes were introduced in the knowledge graph as new nodes representing the pair, and subsequently treated as nodes by the GNNs (see **section “GNN architectural adaptations to classify pairs-of-nodes as new nodes”**). Such node classification tasks involved synthetic lethality (SL) and synthetic non-lethality (SNL) prediction, both for human and yeast, independently.

In the case of yeast, SL and SNL pairs in yeast were obtained from Costanzo et al. (39), where ∼23 million double mutants from Saccharomyces Cerevisiae were evaluated (**Methods**). For the purpose of our study, SL pairs were defined at varying thresholds of double mutant fitness (DMF) progressively corresponding from a lesser to a more stringent definition: DMF <0.5, <0.4, <0.3; respectively corresponding to 290,352, 164,260 and 72,718 pairs). Importantly for our work, only non-essential genes were involved in either SL or SNL pairs. Furthermore, all SNL pairs were composed of genes that were also participating in SL pairs, except for the gene *PTK1*, which was only found in SNL pairs. When training PAGAN on balanced subsets of SL and SNL pairs (**Methods**), models led to good predictive performances on the corresponding train and left out test sets of gene pairs, both in terms of AUROC and AUPR (**Figure 2, Supplementary Figure 2 and Supplementary Table 2**). Performances were higher when using stricter thresholds of DMF values. The PAGAN approach actually led to competitive, yet inferior performances than the ones obtained through the corresponding GNN approach for an edge prediction task trained on identical subsets. This is expected as, as indicated above, in edge prediction tasks, the prediction for each edge used the neighborhood’s information as well as the nodes’ own features connected by the edge.

**Figure 2.**
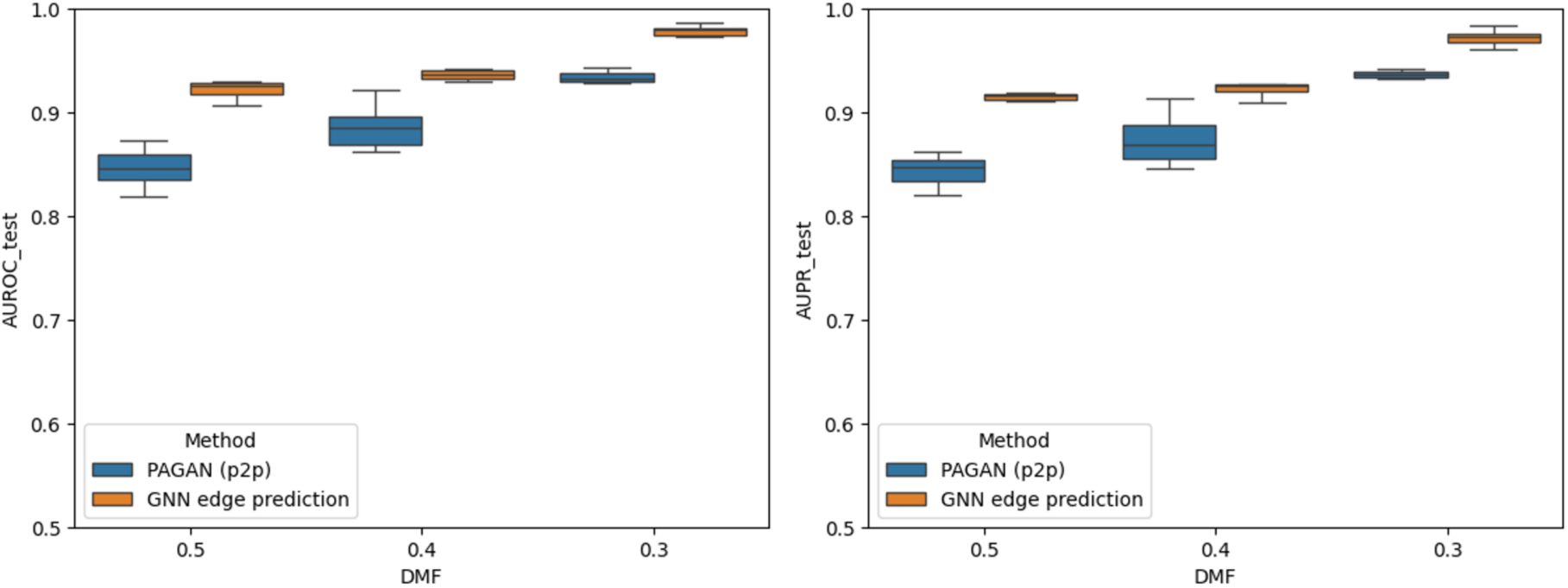
Test performance of the PAGAN approach in the pairs-to-pairs (p2p) setting for synthetic lethality prediction in yeast. Test performances are shown for the classification of Synthetic Lethal (SL) versus Synthetic Non-Lethal (SNL) gene pairs using balanced test sets. The Area Under the Receiver Operating Characteristic curve (AUROC; **left panel**) and the Area Under the Precision-Recall curve (AUPR; **right panel**) are reported for SL pairs defined using increasingly stringent thresholds of double-mutant fitness (DMF): DMF < 0.5, DMF < 0.4, and DMF < 0.3. Results are presented for the PAGAN approach applied in the pairs-to-pairs setting (blue; labelled PAGAN p2p) as well as for the corresponding Graph Neural Network (GNN) edge-prediction approach trained on identical data subsets (orange). Performance values are displayed as boxplots representing the distribution across the six hyperparameter combinations evaluated (**Methods**). Additional details, including the total numbers of SL and SNL pairs included in the training and test sets for each subset, are provided in **Supplementary Table 2**.

In the case of human, SL pairs were obtained from the SyntheticLethalDatabase (Methods) (30,31). For the purpose of the pairs-to-pairs (p2p) approach, SL pairs specific of the myelogenous leukemia cell line K562 were considered, as it was the only cell line with a representative number of SL pairs to perform supervised learning in this setting (Supplementary Table 3). Only synthetic lethal pairs proven *in vitro* with CRISPR or RNAi experiments, were retained. Furthermore, we filtered out SL pairs that were composed of at least one gene considered as “essential” in the K562 cell line. Human SNL pairs were generated by randomly sampling non-essential genes by using two different strategies, referred as matched and unmatched, depending on whether one gene was drawn from the pool of non-essential genes involved in SL pairs and the other from non-essential genes not found in SL pairs (matched), or whether both genes were drawn exclusively from non-essential genes not found in SL pairs (unmatched; **Methods**).

When training PAGAN on balanced subsets of SL and SNL pairs (**Methods**), models led again to good predictive performances on the corresponding train and left out test sets of gene pairs, both in terms of AUROC and AUPR (**Figure 3, Supplementary Figure 3 and Supplementary Table 3**). Performances were generally higher under the unmatched settings. The PAGAN approach actually led to competitive performances to those obtained through the corresponding GNN approach for an edge prediction task trained on identical subsets. PAGAN ranked especially high when considering SL pairs proven *in vitro* based on CRISPR experiments. In addition, PAGAN consistently outperformed SLGNN (**Figure 3**), a GNN–based method that was previously reported to outperform alternative state-of-the-art methods for SL prediction in human, and was thus considered a baseline in our study (49).

**Figure 3.**
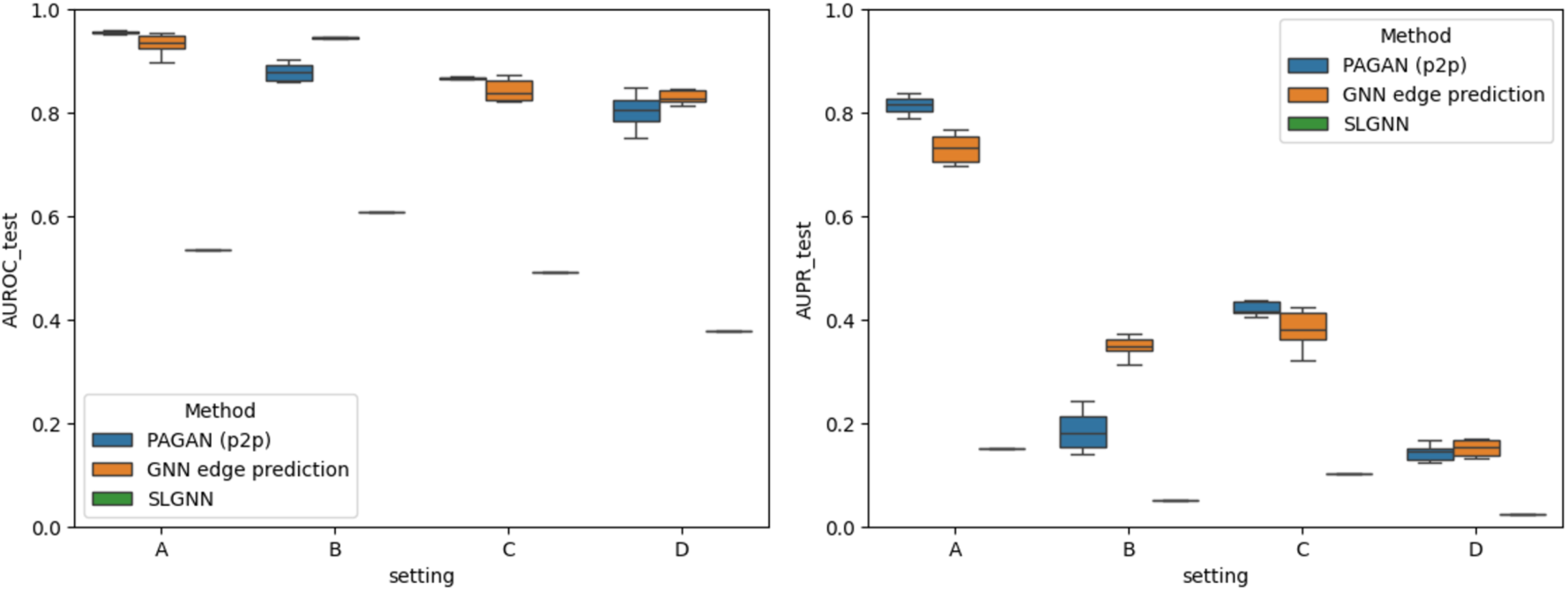
Test performance of the PAGAN approach in the pairs-to-pairs setting for synthetic lethality prediction in human. Test performances are shown for the classification of Synthetic Lethal (SL) versus Synthetic Non-Lethal (SNL) gene pairs using balanced test sets. The Area Under the Receiver Operating Characteristic curve (AUROC; **left panel**) and the Area Under the Precision–Recall curve (AUPR; **right panel**) are reported. Results are shown for the PAGAN approach applied in the pairs-to-pairs setting (blue; labeled PAGAN p2p) and for the corresponding Graph Neural Network (GNN) edge-prediction approach trained on identical subsets (orange). Performance values are displayed as boxplots representing the distribution across the six hyperparameter combinations evaluated (**Methods**). In addition, performances are also reported for SLGNN, a GNN–based method representative of the state-of-the-art methods for SL prediction in human, and considered here as a baseline. The x-axis denotes the different experimental settings defined by the strategy used to generate human SNL pairs through random sampling of non-essential genes. Settings **A** and **B** correspond to the unmatched strategy (i.e. in each SNL pair, one gene was sampled from the pool of non-essential genes involved in SL pairs and the other from non-essential genes not found in SL pairs), whereas settings **C** and **D** correspond to the matched strategy (i.e. both genes in each SNL pair were sampled exclusively from non-essential genes not found in SL pairs). In addition, settings **A** and **C** include SL pairs validated in vitro by either CRISPR or RNAi experiments, while settings **B** and **D** include only SL pairs supported by CRISPR experiments (**Methods**). Further details, including the total numbers of SL and SNL pairs in the training and test sets for each subset, are provided in **Supplementary Table 3**.

Considered together, the results obtained both on yeast and human showed PAGAN’s capacity to learn through GNNs biological network characteristics associated to synthetic lethal gene pairs by representing them as new nodes in the knowledge graph. Such strategy led to competitive performances as compared to the corresponding GNN approach for an edge prediction task, where the goal is to predict node connections in the knowledge graph. It is worth noting that under such edge-prediction task setting, the final prediction for each edge is computed using the neighborhood’s information as well as the nodes’ own features connected by the edge. This is in contrast to the PAGAN approach, where the genes’ own features composing the SL and SNL pairs were not considered, thus supporting the possibility of learning and predicting gene-pairs characteristics without considering their composing gene features.

#### 2.3. Learning on individual genes, predicting on pairs of genes: the *genes-to-pairs* setting

As previously motivated, we hypothesized that pairs of genes leading to synthetic lethality would present, when jointly considered, biological network characteristics similar to essential genes. To address this hypothesis, we evaluated the PAGAN approach by its capacity to learn biological network characteristics associated to gene essentiality and generalize them to the identification of synthetic lethal gene pairs.

In the case of yeast, we first trained a PAGAN model to classify the 1311 essential and 5268 non-essential genes in the knowledge-graph. When evaluated on its ability to discriminate SL from SNL gene pairs (**Methods**), the model achieved average AUROC values ranging from 66.68% to 69.36% and average AUPR values from 64.74% to 67.68% (**Figure 4 and Supplementary Table 4**). Performance improved as increasingly stringent definitions of SL pairs were applied, with higher AUROC and AUPR values obtained for stricter double mutant fitness thresholds (DMF < 0.5 to DMF < 0.3). Notably, performance rose further, up to AUROC = 71.07% to 74.25% and AUPR = 69.47% to 71.97%, respectively, when using specific hyperparameter combinations, i.e. embedding dimension = 32 and ReLU activation (**Methods**). Of note, these hyperparameter settings differ from those yielding the highest performance when training and testing the analogous model on essential genes in the genes-to-genes setting described above. This highlights that hyperparameters optimized for gene essentiality prediction do not necessarily translate to optimal performance for synthetic lethality prediction.

**Figure 4.**
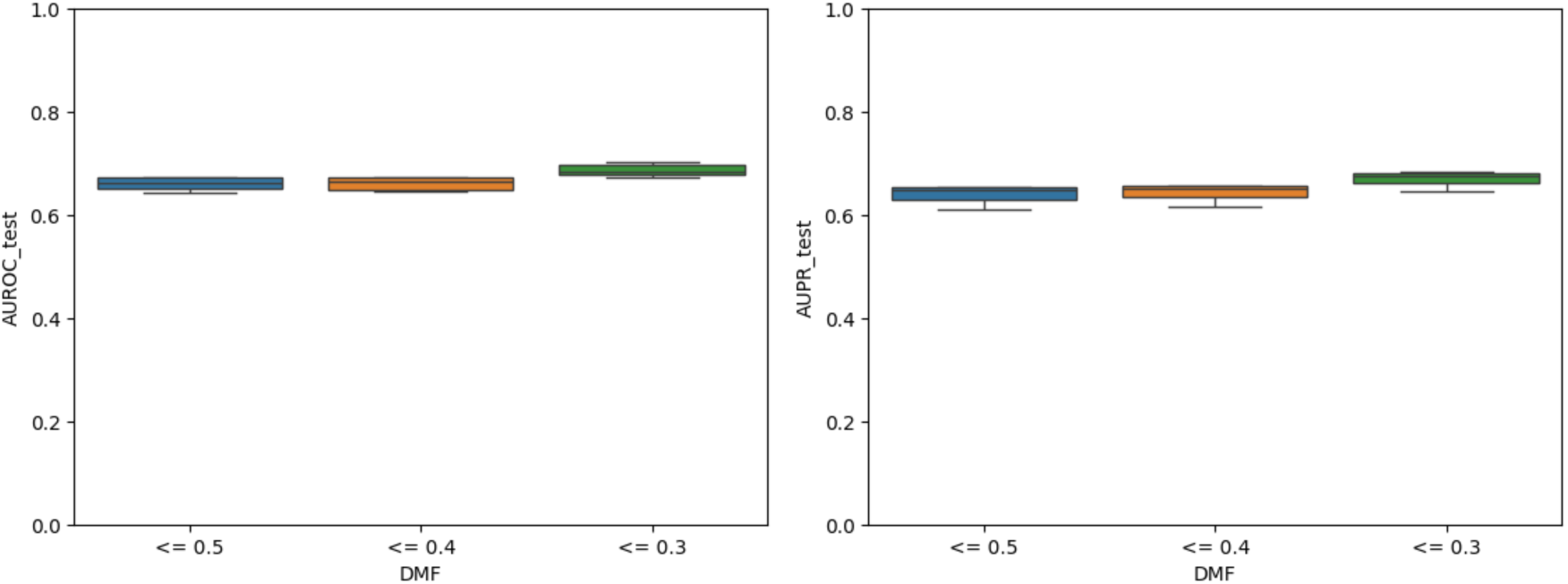
Test performance of the PAGAN approach for synthetic lethality prediction in yeast by a model trained to classify yeast essential genes. Test performances are reported for the classification of Synthetic Lethal (SL) versus Synthetic Non-Lethal (SNL) gene pairs achieved by the PAGAN approach applied in the genes-to-pairs setting. PAGAN models were trained to classify the 1311 essential and 5268 non-essential genes in the yeast knowledge-graph. The Area Under the Receiver Operating Characteristic curve (AUROC; **left panel**) and the Area Under the Precision-Recall curve (AUPR; **right panel**) are reported for SL pairs defined using increasingly stringent thresholds of double-mutant fitness (DMF): DMF < 0.5, DMF < 0.4, and DMF < 0.3. Performance values are displayed as boxplots representing the distribution across the six hyperparameter combinations evaluated (**Methods**). Additional details are provided in **Supplementary Table 4**.

In the case of human, we trained three separate cell-line specific GNN models to classify genes according gene essentiality/non-essentiality, by using the corresponding lists of essential and non-essential genes established by CRISPR on the K562, A375 and A549 cell-lines, respectively. Accordingly, we evaluated them by its ability to discriminate the corresponding cell-line specific lists of SL and SNL pairs, where we considered either all experimentally proven *in vitro* SL pairs, or only those established through CRISPR experiments (**Figure 5**). Of note, among the cell line-specific SL pairs, we filtered out the pairs of genes that were composed of at least one gene considered as “essential” according to strain-specific essential lists (**Methods**). The results showed a consistent capacity to discriminate across the three cell lines SL pairs established through CRISPR experiments, with average AUROC values of 81.37±0.88 %, 94.36±2.08 % and 70.23±1.18 %, for K562, A375 and A549, respectively. More inconsistent results across cell lines were however obtained when considering less confident SL pairs including interfering-RNA experimental protocols (**Supplementary Table 5**).

**Figure 5.**
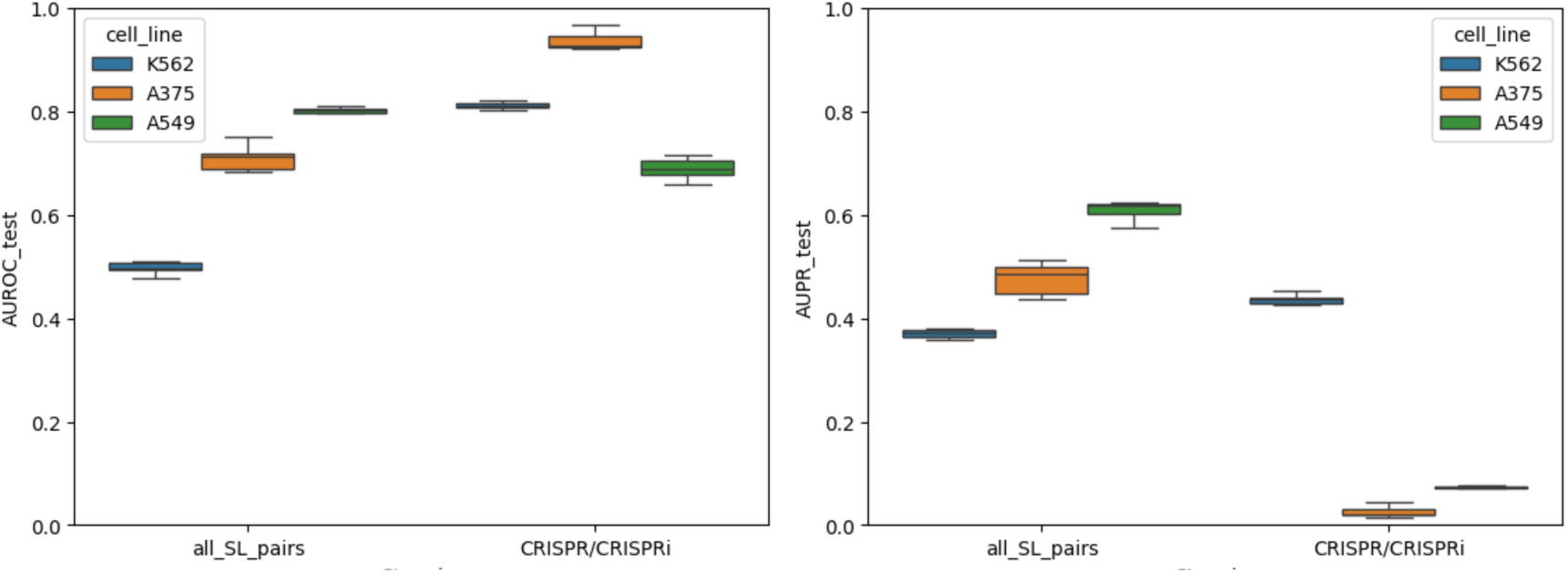
Test performance of the PAGAN approach for cell-line specific synthetic lethality prediction in human cell lines by models trained to classify cell-line specific essential genes. Test performances are reported for the classification of Synthetic Lethal (SL) versus Synthetic Non-Lethal (SNL) gene pairs achieved by the PAGAN approach applied in the genes-to-pairs setting. The performances are shown for three separate cell-line specific PAGAN models trained to classify genes according gene essentiality/non-essentiality, by using the corresponding lists of essential and non-essential genes on the K562 (blue), A375 (orange) and A549 (green) cell-lines. Performances thus correspond to its ability to discriminate the corresponding cell-line specific lists of SL and SNL pairs, where we considered either all experimentally proven *in vitro* SL pairs (denoted in the x-axis as “all_SL_pairs”), or only those established through CRISPR experiments. The Area Under the Receiver Operating Characteristic curve (AUROC; **left panel**) and the Area Under the Precision–Recall curve (AUPR; **right panel**) are reported as boxplots representing the distribution across the six hyperparameter combinations evaluated (**Methods**). Further details, including the total numbers of SL and SNL pairs in the training and test sets for each subset, are provided in **Supplementary Table 5**.

In the previous analysis, similar performances were obtained, both in yeast and human, when excluding from the training set the non-essential genes participating in SL or SNL pairs, i.e. minimizing the risk of data leakage between train and test sets under the genes-to-pairs setting (**Supplementary Tables 4 and 5**). Considered together, these results support the ability of the PAGAN approach to learn biological network characteristics associated with gene essentiality and to generalize this knowledge to the identification of synthetic lethal and non-lethal gene pairs, both of which, by definition, consist exclusively of non-essential genes. This generalization is particularly notable in yeast, where all SNL pairs were composed of genes that also participated in SL pairs (with the exception of PTK1, which was found only in SNL pairs). These observations further support that PAGAN’s discriminative capacity is not merely driven by the individual genes within each pair, but instead reflects network-level properties that, although learned from individual genes, can be effectively applied to pairs of genes.

### 3. Mendelian monogenic disease genes and digenic disease pairs

We applied the PAGAN approach to the task of discriminating n = 4,541 Monogenic Mendelian Disease Genes (MMDGs) from n= 14362 non-disease genes using the human knowledge graph (**Methods**). To assess its ability to identify MMDGs not seen during training, we divided all genes into 10 genomic partitions, stratified to preserve the proportion of MMDGs and non-disease genes in each group. Ten GNN models were then trained independently, each using nine partitions for training and leaving out the remaining partition for testing. This strategy allowed every gene to be scored by a model that had not been trained on it. In this experimental setting, PAGAN achieved a mean AUROC of 77.41% (±0.60) and a mean AUPR of 49.14% (±1.11), with the best-performing model, using a *tanh* activation and embedding dimension 16, reaching an AUROC of 77.89% and an AUPR of 50.10%. These results further support PAGAN’s capacity to learn individual gene characteristics from MMDG that can be generalized to the identification of novel candidate genes, consistent with the results obtained on the study of gene essentiality.

We then retrained the PAGAN approach on the same task of discriminating n=4,541 Monogenic Mendelian Disease Genes (MMDGs) from n=14,362 non-disease genes, as described above, but this time using all genes simultaneously within a single GNN model (**Methods**). We evaluated this model on its ability to discriminate a curated set of n=255 unique human disease gene pairs from n=204,583 unique non-disease gene pairs derived from the Ogloblinsky benchmark dataset (12), which is based on the OLIDA database (6,7). In this setting, the models achieved a mean AUROC of 78.46± 1.55, and a mean AUPR of 0.80%± 0.25%, with the best-performing configurations reaching an AUROC of 81.09% and an AUPR of 0.84% when using a ‘relu’ activation function and an embedding dimension of 32 or reaching an AUROC of 79.53% and an AUPR of 1.27% when using a ‘relu’ activation function and an embedding dimension of 16.

Although these results are positive, their value is limited by the composition of the gene pairs in this benchmark. Specifically, among the 255 disease-associated gene pairs, 197 pairs consisted of two MMDGs, and an additional 50 pairs included one MMDG. As a result, this dataset does not allow us to determine whether PAGAN’s ability to distinguish disease from non-disease gene pairs primarily reflects the signal carried by the MMDGs composing the pairs’ genes, on which the model was explicitly trained.

Furthermore, we benchmarked the previous PAGAN model on a subset of the Ogloblinsky benchmark dataset consisting of n = 69 non-unique disease gene pairs and n = 51,038 non-unique non-disease gene pairs, which had previously been used as an external test set for several state-of-the-art supervised methods for disease gene pair prediction, including DiGePred (8), DIEP (9), Varcopp2.0 (10) and ARBOCK (11) (**Methods**). These methods were trained on a separate set of n = 316 non-unique disease gene pairs and n = 157,292 non-unique non-disease gene pairs, respectively. **Figure 6** presents the ROC and precision-recall (PR) curves obtained for each evaluated method on this test set. Consistent with the results observed on the full set of unique gene pairs from the Ogloblinsky benchmark dataset, PAGAN was able to discriminate disease from non-disease gene pairs, although its performance was inferior to that of methods explicitly trained in a supervised manner on gene pairs. The conclusions drawn from this benchmark are, however, again limited by the composition of the training and test sets: while no significant overlap was detected between disease and non-disease gene pairs across the training and test splits, a substantial overlap was observed in terms of the individual genes composing these pairs (**Supplementary Figure 4**).

**Figure 6.**
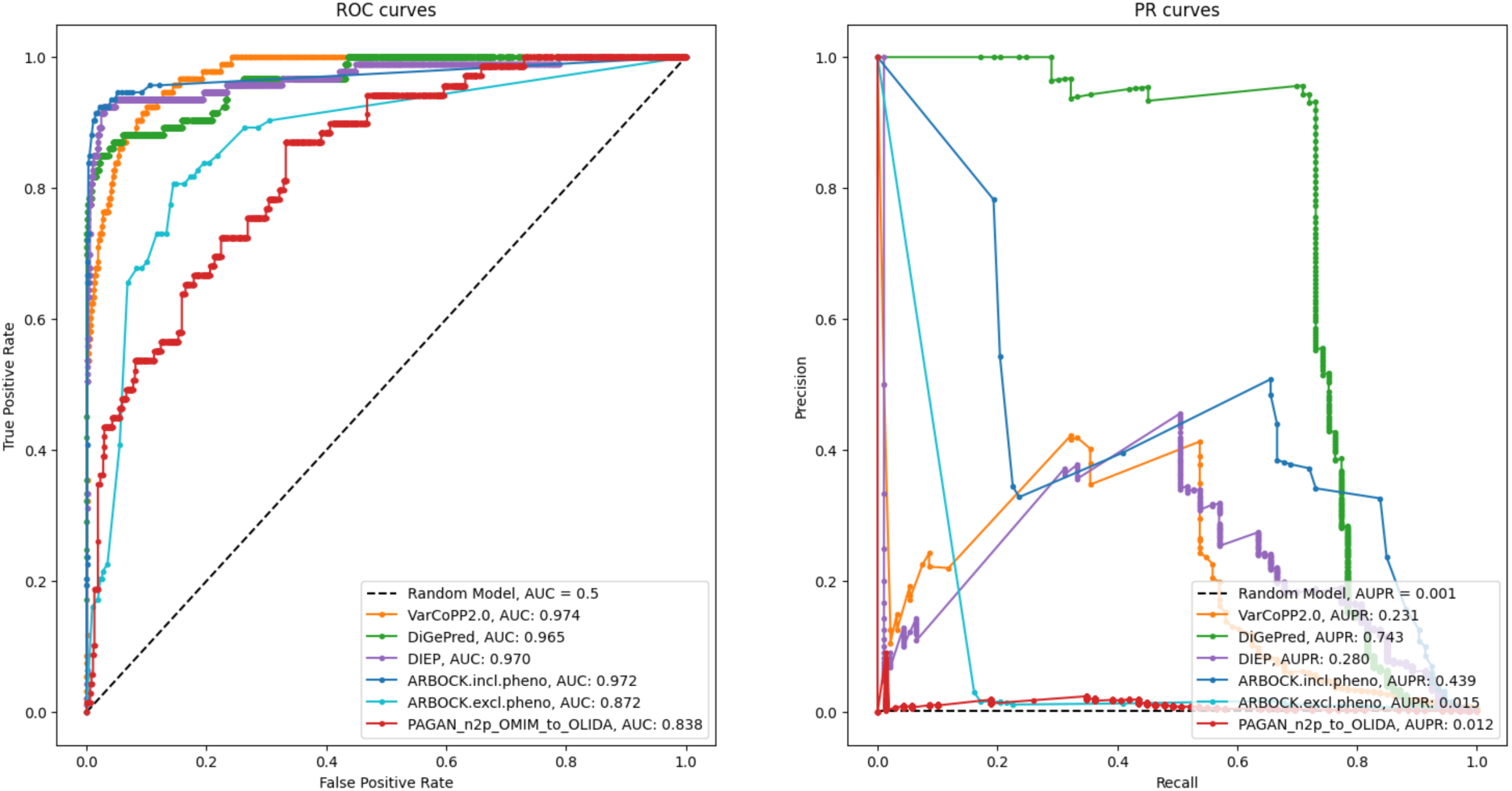
Benchmarking of the PAGAN genes-to-pairs approach against state-of-the-art methods for digenic disease gene pair prediction on the Ogloblinsky benchmark test set. Receiver Operating Characteristic (ROC; left panel) and Precision–Recall (PR; right panel) curves are shown for the discrimination of disease-associated versus non-disease gene pairs on a benchmark test set consisting of n = 69 non-unique disease gene pairs and n = 51,038 non-unique non-disease gene pairs, previously used as an external test set for supervised digenic disease prediction methods (Methods). Curves are reported for PAGAN, applied in the genes-to-pairs setting using a model trained to discriminate Monogenic Mendelian Disease Genes (MMDGs) from non-disease genes (PAGAN g2p, OMIM-to-OLIDA,), as well as for VarCoPP2.0, DiGePred, DIEP, and ARBOCK, these later evaluated either using or not phenotypic information as input). The supervised methods were trained on a separate dataset comprising n = 316 non-unique disease gene pairs and n = 157,292 non-unique non-disease gene pairs. The dashed diagonal line indicates random performance. The corresponding AUROC and AUPR values for each method are reported in the legend of each panel. In the figure, the PAGAN g2p, OMIM-to-OLIDA results corresponds to the model trained with hyperparameter configuration using ‘relu’ activation function and an embedding dimension of 32. Across the six hyperparameter configurations, the mean and standard deviation values were AUROC = 82.587±2.481 % and AUPR = 1.257±0.675%.

Taken together, these results indicate that PAGAN can learn biological network characteristics from Monogenic Mendelian Disease Genes and generalize them to the prediction of digenic disease gene pairs; however, the interpretability of these findings is constrained by the current limited knowledge of digenic disease, as most reported disease-associated pairs are composed predominantly of MMDGs. While PAGAN underperformed alternative methods explicitly trained in a supervised manner on gene pairs, the evaluation of such methods may be biased by the small number of available disease-associated pairs (255 pairs collectively involving only 342 genes) and by substantial overlap between training and test sets in terms of the genes composing the pairs. In contrast, PAGAN is not inherently constrained by the limited size of digenic training datasets, as it is trained on individual MMDGs and subsequently applied to score gene pairs.

### 4. Mendelian monogenic disease genes, complex disease genes and pairs of complex disease genes

The previous evaluation of PAGAN’s ability to learn features of Monogenic Mendelian Disease Genes (MMDGs) and generalize them to the prediction of digenic disease gene pairs was constrained by the limited current knowledge of such pairs, which are predominantly composed of a small subset of MMDGs. To further evaluate such capacity from a complementary angle, we hypothesized that Complex Mendelian Disease Genes (CMDGs) may represent a class of genes likely to be overrepresented in potential digenic disease mechanisms, as they are known to be associated with disease phenotypes but do not follow a strictly monogenic Mendelian mode of inheritance. We thus obtained a representative set of CMDGs from the OMIM database, defined as genes with known molecular disease basis (i.e. a ClinVar supporting evidence level of 3), not flagged as somatic, and associated with phenotypes whose descriptions contained indicators of complex genetic architectures, reflecting a non-monogenic contribution to Mendelian disease (**Methods**).

We then investigated how the previously described PAGAN approach, trained to discriminate n = 4,541 MMDGs from n = 14,362 non-disease genes, scored a disjoint independent set of n = 324 CMDGs, as well as their pairwise combinations (n = 52326 gene pairs). To this aim we used the ten GNN models trained independently, each using nine genomic partitions for training and leaving out the remaining partition for testing, allowing every gene to be scored by a model that had not been trained on it. Importantly for this analysis, the list of CMDG was not part of the corresponding PAGAN training set across any of such models. Results showed that CMDGs exhibited PAGAN scores intermediate between MMDG and non-disease genes (**Figure 7**). Interestingly, PAGAN scores for CMDG inversely associated with the degree of disease specificity of genes, as measured by the gene’s Disease Specificity Index (DSI) and the gene’s Disease Pleiotropy Index (**Supplementary Figure 5**; **Methods**). Thus, CMDGs with the more pathogenic PAGAN predictions presented broader disease associations (i.e. lower DSI) and associated with more diverse disease classes (i.e. higher DPI values). Of note, such associations were not a direct correlate of the node centrality of a gene in the underlying protein-protein interaction network (**Supplementary Figure 5**). Finally, the distribution of PAGAN pathogenicity scores of the gene pairs composed of two CMDG (n = 52326) closely resembles that of MMDG, and was significantly higher than those of gene pairs composed of one CMDG and one non-disease gene (two-sided Wilcoxon rank sum test p-value = 0.00e+00).

**Figure 7.**
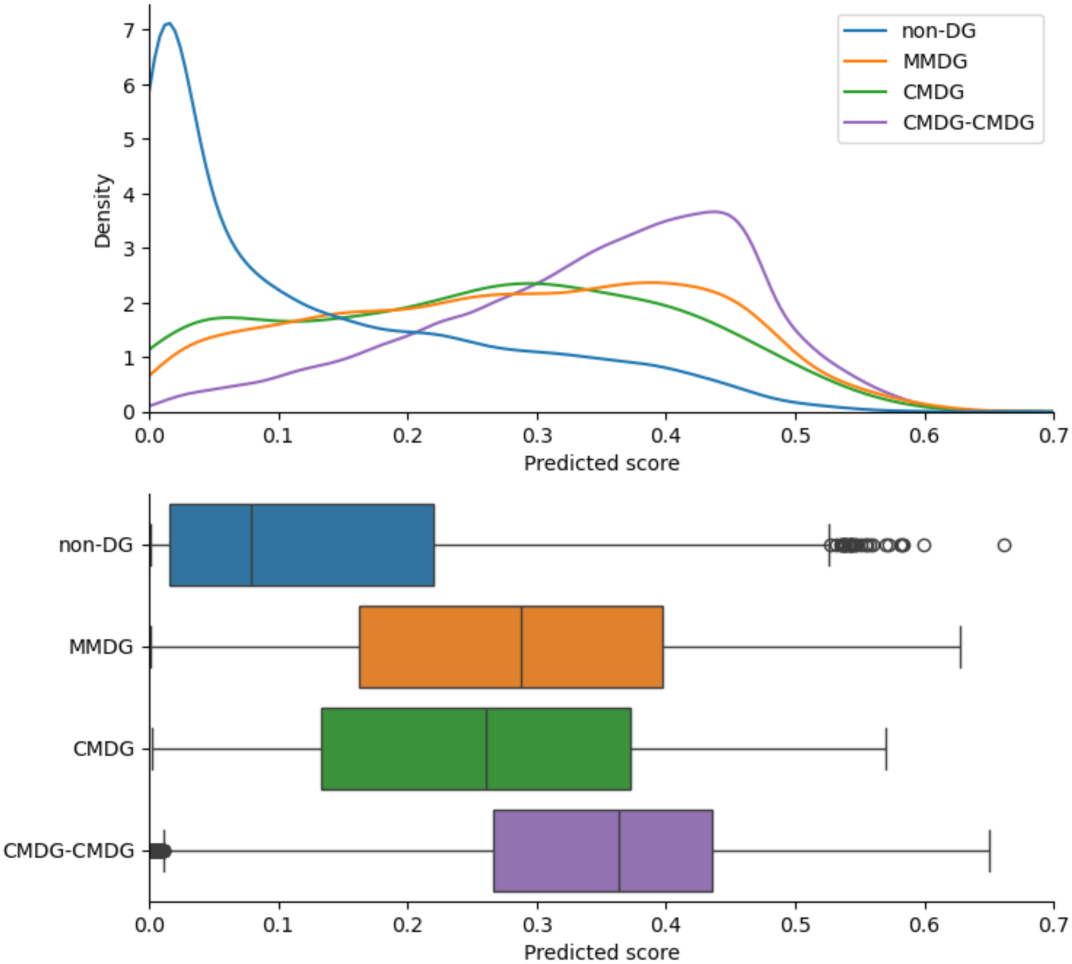
Distribution of PAGAN pathogenicity scores for non-disease genes, monogenic Mendelian disease genes, complex Mendelian disease genes, and their pairwise combinations. The figure shows the distribution of PAGAN pathogenicity scores obtained from a model trained to discriminate n = 4,541 Monogenic Mendelian Disease Genes (MMDGs) from n = 14,362 non-disease genes. Scores are reported for non-disease genes, monogenic OMIM disease genes, an independent and disjoint set of n = 324 Complex Mendelian Disease Genes (CMDGs), and all pairwise combinations of CMDGs (n = 52,326 gene pairs). The top panel displays density estimates of the score distributions, while the bottom panel summarizes the same distributions using boxplots. For this analysis, PAGAN scores were obtained using ten independently trained GNN models based on genomic partitioning, such that each gene was scored by a model that did not include it in the training set. Importantly, CMDGs were not used during model training, allowing an unbiased assessment of how PAGAN generalizes from monogenic disease genes to complex disease genes and their combinations.

The previous results suggest that the biological network properties associated with Monogenic Mendelian Disease Genes and learned by the PAGAN approach are partially recapitulated in Complex Mendelian Disease Genes. Specifically, the more similar the network context of a CMDG is to that of MMDGs, the more susceptible it is to participate in complex disease genetic architectures in diverse combinations with other genes, ultimately associating to a larger spectrum of phenotypic consequences. Furthermore, digenic combinations of CMDG generally presented PAGAN pathogenicity scores comparable to those observed for Monogenic Mendelian Disease genes, suggesting the possibility that such digenic combinations of CMDG could be a substrate of human disease phenotypes as severe as those caused by MMDG.

## Discussion

In this study, we introduced and evaluated PAGAN, a graph representation learning framework designed to infer properties of gene pairs by learning from phenotypically-analogous single-gene characteristics in heterogeneous biological networks. Using multiplex knowledge graphs from yeast and human, we showed that PAGAN can effectively capture biological network characteristics associated with gene essentiality and Monogenic Mendelian disease genes, and extrapolate them, respectively, to the identification of synthetic lethal and digenic disease gene pairs. In a genes-to-genes setting, PAGAN demonstrated that GNN-based architectures outperform traditional machine learning approaches for gene essentiality prediction, while a modified architecture that excludes gene-intrinsic features retained comparable performance and enabled applicability to featureless nodes. Extending this framework to pairs of genes, we showed that representing gene pairs as new nodes allows PAGAN to learn and predict synthetic lethal interactions competitively with dedicated edge-prediction approaches, which represent the current state-of-the-art (51), despite relying exclusively on network context rather than gene-level features. Finally, PAGAN was able to generalize knowledge learned from individual genes to gene pairs, supporting the hypothesis that synthetic lethal pairs share network-level properties with essential genes, and that digenic disease pairs partially recapitulate properties of monogenic Mendelian disease genes. Together, these results establish PAGAN as a unified and flexible approach for transferring biological network signatures learned at the gene level to digenic interactions, without requiring large curated datasets of labeled gene pairs.

While PAGAN achieved positive results on the Ogloblinsky benchmark dataset (12), its performance was on average lower than that of current state-of-the-art methods for prediction of pathogenic digenic interactions, which are based on supervised learning on gene pairs. Direct comparison should, however, be interpreted with caution, as these approaches address partially different prediction tasks. In particular, VarCoPP2.0 operates at the level of variant pairs, whereas DIEP, DiGePred, ARBOCK, and PAGAN aim to predict disease involvement at the gene-pair level, thus assigning identical scores to all variant combinations affecting the same pair of genes. More importantly, several characteristics of the OLIDA database and of supervised models trained on it need some consideration. While OLIDA constitutes a key resource for reported cases of digenic and oligogenic inheritance, most variant pairs lack functional validation and are primarily supported by segregation analyses and phenotype-based enrichment. Consequently, many reported pairs involve at least one gene already implicated in monogenic disease, blurring the distinction between strictly digenic mechanisms and modifier effects. This is reflected in the frequent classification of cases as involving one major gene and one modifier gene, and in the relatively small number of pairs reported as unequivocally digenic. Finally, current reference methods such DiGePred, DIEP, VarCopp2.0 and ARBOCK, rely on gene–phenotype associations, particularly on Human Phenotype Ontology annotations, which are often central to the initial reporting of candidate digenic pairs. The dependence on gene-level HPO annotations as gene features during the supervised learning process may introduce biases and contribute to overfitting, as suggested by the reduced performance of ARBOCK when phenotype information is excluded. Together, these limitations highlight the challenges inherent to benchmarking digenic prediction methods and motivate complementary approaches that do not rely on supervised learning on the limited and probably biased set of currently known digenic interactions in human disease.

The ability of GNN-based approaches to learn disease-relevant features from biological knowledge graphs, as demonstrated in this work, is consistent with recent advances in network medicine that increasingly rely on graph representation learning (52–54). The current implementation of PAGAN was designed as a proof of concept to evaluate the capacity of GNNs to generalize gene-level characteristics to phenotypically analogous properties arising from digenic interactions. From this perspective, we deliberately explored a range of hyperparameter settings rather than performing task-specific hyperparameter optimization for single gene classification, as such optimization may not be optimal for generalization to gene-pair classification and could potentially attenuate the relevant predictive signal. Hyperparameter tuning specifically tailored to the genes-to-pairs setting therefore remains an open area for future investigation. In particular, in this work we limited the exploration of the depth of GNN architectures to at most three graph convolutional layers, corresponding to a receptive field extending up to three hops in the knowledge graph. While deeper architectures could in principle capture longer-range dependencies, increasing the number of layers in GNNs is known to introduce over-smoothing (i.e. where node representations become increasingly similar), and over-squashing (i.e. excessive information compression through bottleneck edges), particularly in small-world biological networks (55). Similarly, alternative GNN formulations oriented toward subgraph classification, in which gene pairs are represented as subgraphs rather than as individual nodes, represent a promising direction for further exploration (56,57). Finally, extending the knowledge graph to incorporate additional gene features and biological relationships, including gene regulatory interactions, post-translational modifications, and signaling pathways, may further enhance the expressiveness and predictive power of the approach.

From a clinical diagnostic perspective, PAGAN scores derived from models trained to classify monogenic Mendelian disease genes may support the identification of candidate digenic disease mechanisms in genetically unresolved rare disease cases. By providing a pre-computed score for all possible protein coding gene pairs, PAGAN enables a systematic prioritization of potential combinatorial gene effects that are not captured by current single-gene analyses pipelines. Particularly interesting is the observation that pairs of complex Mendelian disease genes exhibited pathogenicity score distributions comparable to those of monogenic disease genes, supporting the hypothesis that such genes may constitute a substrate for rare disease phenotypes arising from digenic perturbations. This may be particularly relevant in the context of whole-genome sequencing (WGS) of trios from sporadic cases in which both parents are asymptomatic and no clearly pathogenic monogenic cause is identified, a situation that is especially frequent in intellectual disability and neurodevelopmental disorder cohorts. In such situations, a plausible hypothesis is that the affected individual carries rare deleterious variants affecting two genes whose combined perturbation leads to disease, despite each variant being insufficient to produce a phenotype on its own. These configurations may include recessive-like digenic inheritance, combined heterozygous loss-of-function effects in haploinsufficient genes, or interactions involving dominant-negative mechanisms. In this context, PAGAN can also be trained on alternative gene-level properties relevant to disease mechanisms, such as haploinsufficiency. The probability of loss-of-function intolerance, or pLI (21), which reflects a gene’s sensitivity to heterozygous truncating variants and is derived from large-scale population data, provides a largely unbiased proxy for this property. Training PAGAN to discriminate haploinsufficient from haplo-tolerant genes based on their pLI value could therefore enable the identification of gene pairs exhibiting elevated joint intolerance to double-heterozygous loss-of-function, even when neither gene would be classified as haploinsufficient on its own. Such combinations may represent an additional class of digenic disease mechanisms that can be highlighted through gene-pair–level prioritization. Finally, interpretation of candidate pairs identified through PAGAN would require further expert assessment of the clinical and biological plausibility of their joint involvement in cellular or physiological processes, and would typically require additional genetic, functional, and experimental studies to establish their contribution to disease.

## Conclusion

In this work we present PAGAN as a graph representation learning framework that infers properties of gene pairs by generalizing phenotypically analogous characteristics learned at the single-gene level from heterogeneous biological networks. Across yeast and human datasets, PAGAN captures network-level features associated with gene essentiality and monogenic Mendelian disease genes and extrapolates them, respectively, to the identification of synthetic lethal and digenic disease gene pairs, without requiring supervised training on large collections of labeled gene pairs. By representing pairs of genes as new nodes embedded in the same biological context as individual genes, PAGAN provides a unified and flexible approach to transfer gene-level information to digenic interactions. While the interpretation of predicted gene pairs requires careful biological and clinical evaluation, this work provides a complementary strategy to explore digenic mechanisms that are not readily addressed by monogenic models and that are difficult to capture using supervised approaches relying on currently reported digenic interactions. By learning from gene-level properties defined on large and well-characterized datasets, PAGAN is not intrinsically constrained by the limited number and restricted diversity of currently known digenic cases, and may therefore facilitate the exploration of a broader spectrum of combinatorial genetic architectures.

## Data Availability

All data used in this study are publicly available and were obtained from the resources described in the Methods section, with corresponding URLs provided therein.

## Abbreviations

ANN: Artificial Neural Network
AUPR: Area Under the Precision–Recall curve
AUROC: Area Under the Receiver Operating Characteristic curve
BP: Biological Process (Gene Ontology)
CC: Cellular Component (Gene Ontology)
CMDG: Complex Mendelian Disease Gene
DMF: Double Mutant Fitness
DIDA: Digenic Diseases Database
DPI: Disease Pleiotropy Index
DSI: Disease Specificity Index
GAT: Graph Attention Network
GCN: Graph Convolutional Network
GDI: Gene Damage Index
GNN: Graph Neural Network
GO: Gene Ontology
HPO: Human Phenotype Ontology
KG: Knowledge Graph
MMDG: Monogenic Mendelian Disease Gene
MF: Molecular Function (Gene Ontology)
MPNN: Message Passing Neural Network
NDD: Neurodevelopmental Disorder
OGEE: Online GEne Essentiality database
OLIDA: OLIgogenic Diseases Database
OMIM: Online Mendelian Inheritance in Man
PAGAN: PAirs of Genes As geNes
PPI: Protein–Protein Interaction
PR: Precision–Recall
RNAi: RNA interference
SL: Synthetic Lethal (or Synthetic Lethality)
SNL: Synthetic Non-Lethal
SNV: Single-Nucleotide Variant
SV: Structural Variant
WGS: Whole-Genome Sequencing

## Declarations

### Ethics approval and consent to participate

Not applicable

### Consent for publication

Not applicable.

### Availability of data and materials

The software developed and used in this study is openly available and will be released upon publication at https://github.com/RausellLab/.

### Competing interests

Authors declare that they have no competing financial and/or non-financial interests, or other interests that might be perceived to influence the results and/or discussion reported in this paper.

### Funding

The Laboratory of Clinical Bioinformatics of the Imagine Institute, headed by A.R. was partly supported by the French National Research Agency (ANR) ‘Investissements d’Avenir’ Program [ANR-10-IAHU-01 and ANR-21-PMRB-0004, FACE.S-4-KIDS project, “FACE and SKULL for Key Innovative Data Science]; by the European Rare Diseases Alliance (ERDERA) program funded by the European Union’s Horizon Europe research and innovation program under grant agreement N°101156595; by the French government as part of the “Important Project of Common European Interest” (IPCEI) Cloud call of the France 2030 program (E2CC - AI4RDP - AI for Rare Dis-eases Pathogenicity project); and by INSERM, AAP 2020 Maladies rares: résoudre les impasses diagnostiques. Projet ResDiCard, resolving diagnostic deadlock in Cardiomyopathies. RN was funded by the MD-PhD program of the Imagine Institute, with the funding contribution of Promepar Asset Management.

### Author contributions

Conceptualization: RN & AR; Data curation: RN; Formal analysis: RN, BC; Funding acquisition: AR; Investigation: RN, BC, VM & AR; Methodology: RN & AR; Project administration: RN & AR; Resources: RN, BC, VM & AR; Software: RN, BC; Supervision: AR, VM; Validation: RN & AR; Visualization: RN,BC; Writing – original draft: RN & AR; Writing – review & editing: RN, BC, VM & AR.

## Acknowledgements

We thank the Bioinformatics platform and IT service of the Institute Imagine for continuous support and to all members of the Clinical Bioinformatics laboratory for constructive feedback.

## Notes

### Competing Interest Statement

The authors have declared no competing interest.

