## Supplementary Material for "PAGAN predicts digenic interactions by generalizing single-gene representations in biological networks"

1. Supplementary Figures
2. Supplementary Tables

### 1. Supplementary Figures

A.

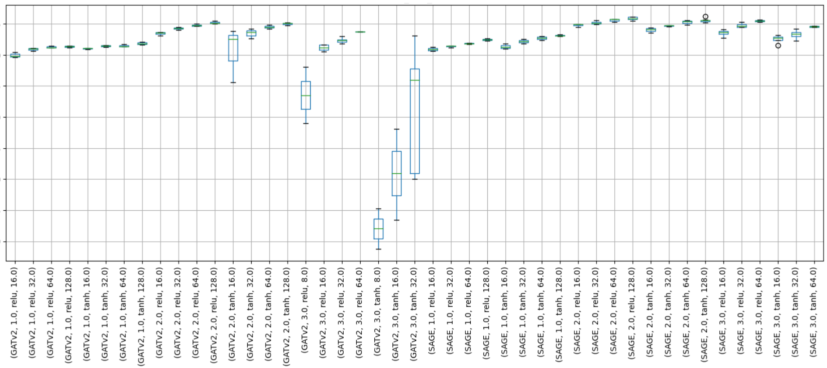


B.

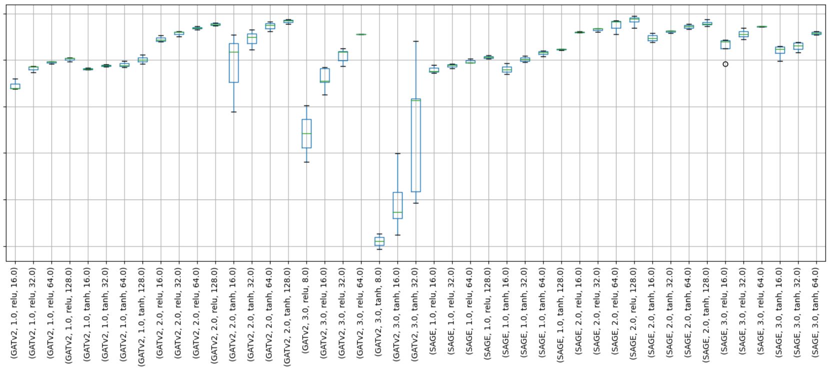


C.

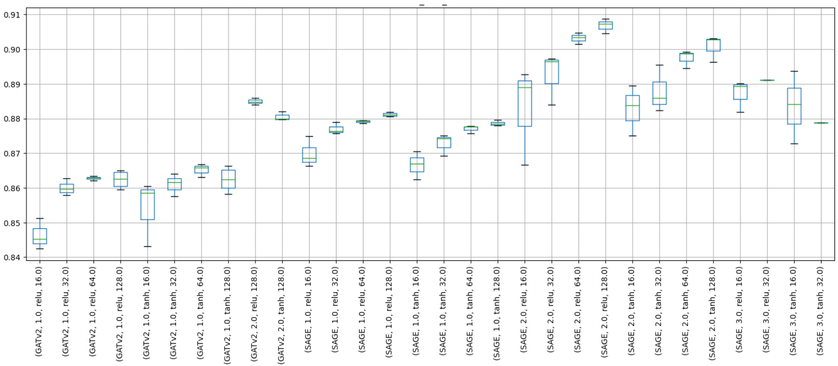


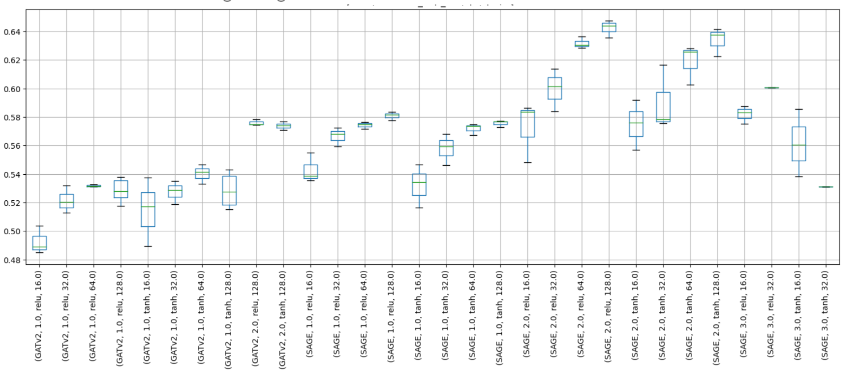


**Supplementary Figure 1.** Hyperparameter exploration of the PAGAN approach in the *genes-to-genes* setting for gene essentiality and non-essentiality prediction. Boxplots represent the distribution of the Area Under the Receiver Operating Characteristic (AUROC, panels **A** and **C**), and the Area Under the Precision–Recall Curve (AUPR, panels B and D) for yeast (panels A and B) and human (panels (**C** and **D**), obtained through 10-fold cross validation on a random train split, representing 70% of the target genes. The combinations of hyperparameters explored are indicated in the x-axis for each boxplot, including: the convolution type (SAGEConv or GATv2Conv), the number of convolutional layers (1, 2, or 3), the activation function (ReLU or tanh) and the batch size (16, 32, 64, or 128).

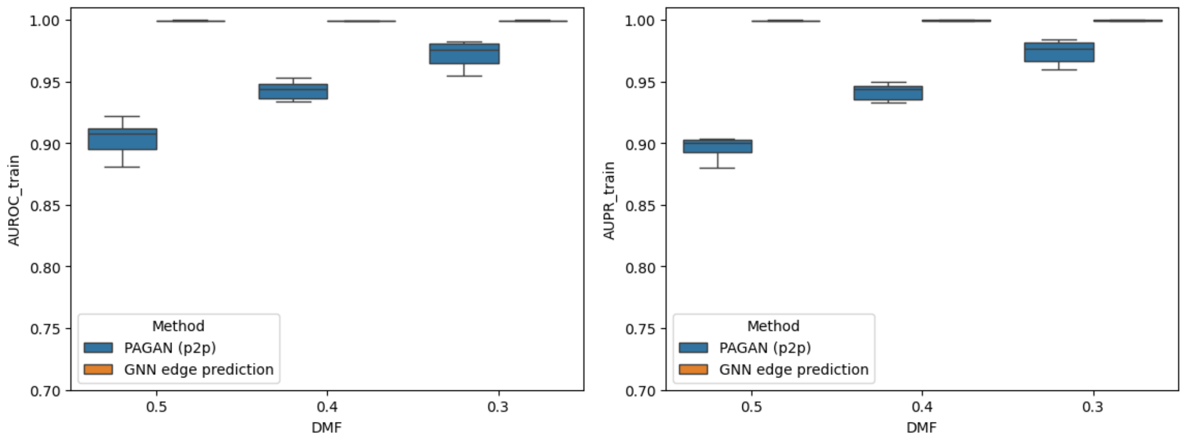


**Supplementary Figure 2: Train performances obtained when training the PAGAN approach on the pairs-to-pairs setting in yeast classifying Synthetic Lethal from Synthetic Non Lethal pairs.** AUROC (**left**) and AUPR (**right**) are reported for the classification of balanced train sets of SL and SNL pairs, where SL pairs were defined at varying thresholds of double mutant fitness (DMF) progressively corresponding from a lesser to a more stringent definition: DMF <0.5, <0.4, <0.3. Results correspond to the PAGAN approach on the pairs-to-pairs setting (colored in blue and indicated as PAGAN p2p) as well as for the corresponding GNN approach for an edge prediction task trained on identical subsets (colored in orange). Performances are reported as boxplots representing the values distribution across the six hyperparameter combinations evaluated (**Methods**). Further details, including the total numbers of SL and SNL pairs composing the train and test of the different subsets are reported in **Supplementary Table 2**.

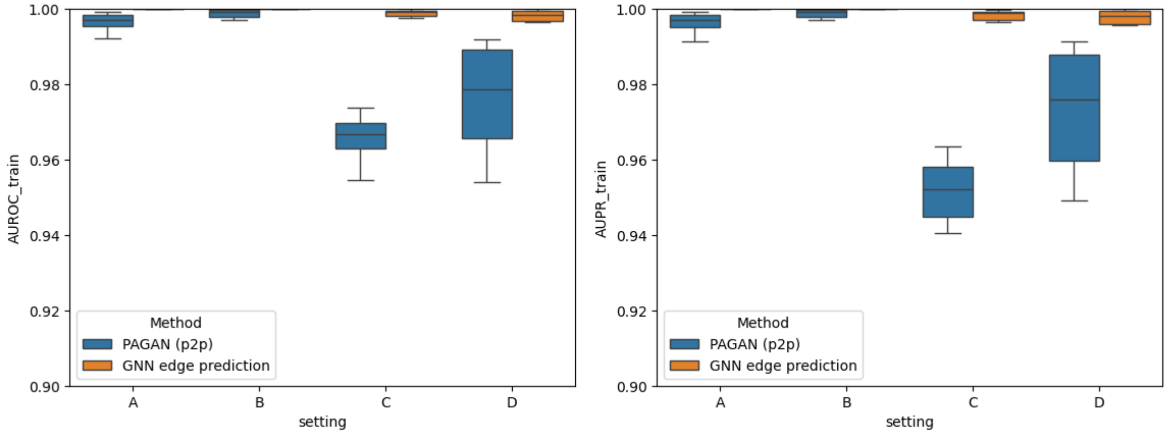


**Supplementary Figure 3. Training performance of the PAGAN approach in the pairs-to-pairs setting for synthetic lethality prediction in human.** Training performances are shown for the classification of Synthetic Lethal (SL) versus Synthetic Non-Lethal (SNL) gene pairs using balanced training sets. The Area Under the Receiver Operating Characteristic curve (AUROC; **left panel**) and the Area Under the Precision–Recall curve (AUPR; **right panel**) are reported. Results are shown for the PAGAN approach applied in the pairs-to-pairs setting (blue; labeled PAGAN p2p) and for the corresponding Graph Neural Network (GNN) edge-prediction approach trained on identical subsets (orange). Performance values are displayed as boxplots representing the distribution across the six hyperparameter combinations evaluated (**Methods**). The x-axis denotes the different experimental settings defined by the strategy used to generate human SNL pairs through random sampling of non-essential genes. Settings **A** and **B** correspond to the unmatched strategy (i.e. in each SNL pair, one gene was sampled from the pool of non-essential genes involved in SL pairs and the other from non-essential genes not found in SL pairs), whereas settings **C** and **D** correspond to the matched strategy (i.e. both genes in each SNL pair were sampled exclusively from non-essential genes not found in SL pairs). In addition, settings **A** and **C** include SL pairs validated in vitro by either CRISPR or RNAi experiments, while settings **B** and **D** include only SL pairs supported by CRISPR experiments (**Methods**). Further details, including the total numbers of SL and SNL pairs in the training and test sets for each subset, are provided in **Supplementary Table 3**.

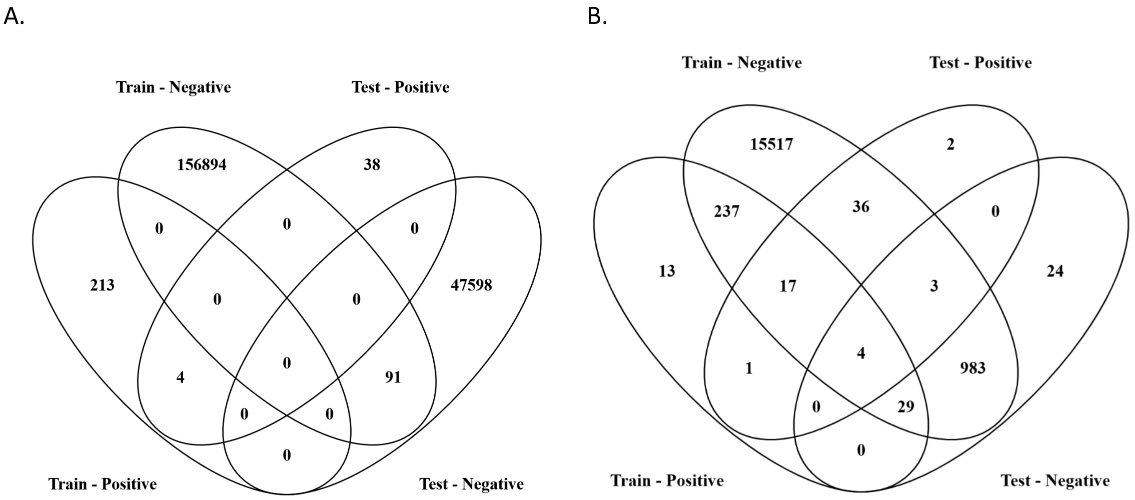


**Supplementary Figure 4.** Venn diagrams representing the intersection beween the Train and Test sets in the Ogloblinsky benchmark dataset. The Ogloblinsky benchmark is composed of curated set of n=255 unique human disease gene pairs from n=204,583 unique non-disease gene pairs derived from the OLIDA database. The total set was used for training and testing supervised learning models for digenic disease prediction (see text). **Panel A** represents the number of unique disease (denoted as Positive) and unique non-disease gene pairs (denoted as Negative) across the training and test sets used in the Ogloblinsky benchmark dataset, and their overlaps. **Panel B** represents the number of unique individual genes composing the Positive and Negative pairs in Panel A, and the overlaps across the training and test sets.

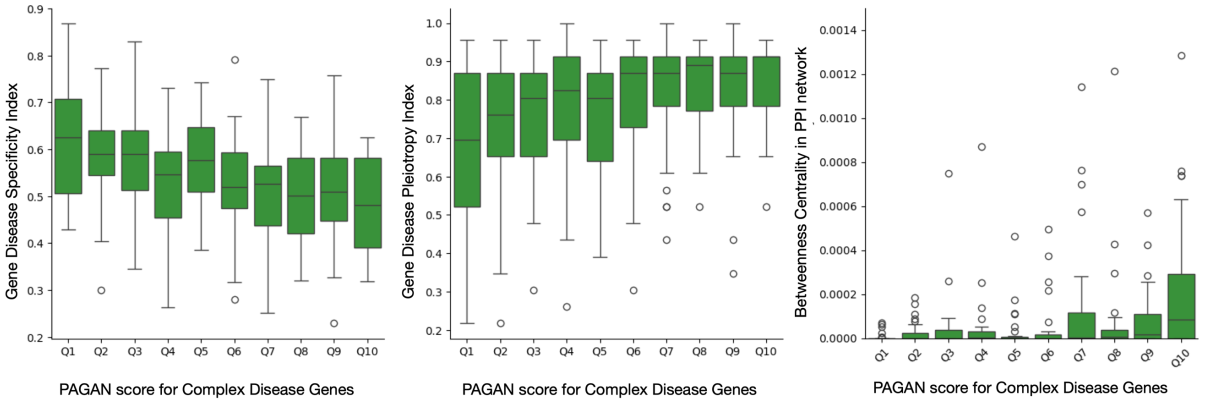


**Supplementary Figure 5. Relationship between PAGAN pathogenicity scores and disease specificity, pleiotropy, and network centrality in Complex Mendelian Disease Genes (CMDGs).** The figure shows the association between PAGAN pathogenicity scores and gene-level properties for a disjoint independent set of n = 325 CMDGs, scored using a PAGAN model trained to discriminate n = 4,541 Monogenic Mendelian Disease Genes (MMDGs) from n = 14,362 non-disease genes. CMDGs were not included in the training set of the model. CMDGs were grouped into bins according to their PAGAN pathogenicity scores (x-axis). Left panel: distribution of the Gene Disease Specificity Index (DSI; y-axis) across PAGAN score bins. Middle panel: distribution of the Disease Pleiotropy Index (DPI; y-axis) across PAGAN score bins. Right panel: distribution of betweenness centrality values in the protein–protein interaction (PPI) network (y-axis) across the same PAGAN score bins. Distributions are shown as boxplots. Spearman rank correlations between PAGAN pathogenicity scores and gene-level properties were as follows: DSI (r = −0.340, p-value = 3.19e-10), DPI (r = 0.306, p-value = 1.85e-08), and PPI betweenness centrality (r = 0.292, p-value = 8.69e-08).

#### 2. Supplementary Tables

| | Yeast | Human |
|---|---|---|
| **nodes** | | |
| GENE | 6579 | 19330 |
| ANATOMY | / | 14337 |
| BP (Biological Process) | 27995 | 27993 |
| CC (Cellular Component) | 4040 | 4039 |
| MF (Molecular Function) | 11297 | 11271 |
| **edges** | | |
| ('GENE', 'PPI', 'GENE') | 1710476 | 1423054 |
| ('GENE', 'paralog', 'GENE') | 11266 | 260780 |
| ('GENE', 'expressed_in', 'ANATOMY') | / | 3135877 |
| ('ANATOMY', 'rev_expressed_in', 'GENE') | / | 3135877 |
| ('ANATOMY', 'ana_ana', 'ANATOMY') | / | 14333 |
| ('ANATOMY', 'rev_ana_ana', 'ANATOMY') | / | 14333 |
| ('GENE', 'gene_to_BP', 'BP') | 23294 | 135647 |
| ('BP', 'rev_gene_to_BP', 'GENE') | 23294 | 135647 |
| ('BP', 'biological_process', 'BP') | 51162 | 51162 |
| ('BP', 'rev_biological_process', 'BP') | 51162 | 51162 |
| ('GENE', 'gene_to_CC', 'CC') | 22222 | 80512 |
| ('CC', 'rev_gene_to_CC', 'GENE') | 22222 | 80512 |
| ('CC', 'cellular_component', 'CC') | 4673 | 4673 |
| ('CC', 'rev_cellular_component', 'CC') | 4673 | 4673 |
| ('GENE', 'gene_to_MF', 'MF') | 17497 | 69943 |
| ('MF', 'rev_gene_to_MF', 'GENE') | 17497 | 69943 |
| ('MF', 'molecular_function', 'MF') | 13833 | 13833 |
| ('MF', 'rev_molecular_function', 'MF') | 13833 | 13833 |

**Supplementary Table 1. Summary of node and edge types in the human and yeast knowledge-graph networks.** The table reports the number of nodes and edges composing the heterogeneous biological knowledge-graph networks for human and yeast. Details of the network construction and data sources are provided in the **Methods** section. Columns correspond to the two species, while rows indicate either node or edge categories. Node types include GENE (protein-coding genes or their encoded proteins), ANATOMY (anatomical entities such as organs, tissues, or cell types, included only for human), and the three Gene Ontology (GO) categories: BP (Biological Process), CC (Cellular Component), and MF (Molecular Function). Edge types represent biological relationships between node types and are denoted as (source node type, relation, target node type). Relations include PPI (protein–protein interaction), paralog (gene–gene paralogy), expressed_in and rev_expressed_in (gene–anatomy expression associations, human only), ana_ana and rev_ana_ana (hierarchical relations among anatomical entities, human only), and gene_to_BP/CC/MF and their reverse edges linking genes to GO terms. Additional relations describe hierarchical links among GO terms within each ontology: biological_process and rev_biological_process, cellular_component and rev_cellular_component, and molecular_function and rev_molecular_function. Bidirectional relations are represented by both the forward and reverse edges.

**A.**

| DMF | PAGAN p2p Average AUROC | PAGAN p2p Max AUROC | GNN edge prediction Average AUROC | GNN edge prediction Max AUROC |
|---|---|---|---|---|
| 0.5 | 84.64±1.95 % | 87.23% (16, 'relu') | 92.22±0.89 % | 93.04% (64, 'tanh') |
| 0.4 | 88.62±2.22 % | 92.16% (16, 'relu') | 93.96±1.14 % | 96.12% (32, 'relu') |
| 0.3 | 93.41±0.61 % | 94.30% (16, 'tanh') | 97.65±1.00 % | 98.64% (32, 'tanh') |

**B.**

| DMF | PAGAN p2p Average AUPR | PAGAN p2p Max AUPR | GNN edge prediction Average AUPR | GNN edge prediction Max AUPR |
|---|---|---|---|---|
| 0.5 | 84.40±1.57 % | 86.23% (16, 'relu') | 91.77±0.74 % | 93.14% (16, 'relu') |
| 0.4 | 87.36±2.56 % | 91.33% (16, 'relu') | 92.58±1.25 % | 94.72% (32, 'relu') |
| 0.3 | 93.65±0.39 % | 94.20% (16, 'tanh') | 97.19±0.79 % | 98.33% (32, 'tanh') |

**C.**

| DMF | SL pairs train set | SNL pairs train set | SL pairs test set | SNL pairs test set |
|---|---|---|---|---|
| 0.5 | 200220 | 200220 | 8048 | 8048 |
| 0.4 | 118779 | 118779 | 3180 | 3180 |
| 0.3 | 49815 | 49815 | 2122 | 2122 |

**Supplementary Table 2. Test performance of the PAGAN approach in the pairs-to-pairs (p2p) setting for synthetic lethality prediction in yeast.** Test performances are reported for the classification of Synthetic Lethal (SL) versus Synthetic Non-Lethal (SNL) gene pairs using balanced test sets. The Area Under the Receiver Operating Characteristic curve (AUROC; **panel A**) and the Area Under the Precision–Recall curve (AUPR; **panel B**) are shown for SL pairs defined using increasingly stringent thresholds of double-mutant fitness (DMF): DMF < 0.5, DMF < 0.4, and DMF < 0.3. Results are presented for the PAGAN approach applied in the pairs-to-pairs setting (denoted PAGAN p2p) as well as for the corresponding Graph Neural Network (GNN) edge-prediction approach trained on identical data subsets. Performance values are reported as the mean and standard deviation across the six hyperparameter combinations evaluated (second and fourth columns in panels A and B; Methods). In addition, the maximum AUROC and AUPR values achieved by any individual hyperparameter configuration are provided (third and fifth columns in panels A and B; **Methods**), together with the corresponding hyperparameters indicated in parentheses, specifying the embedding dimension and activation function ('relu' or 'tanh'). The total numbers of SL and SNL pairs included in the training and test sets for each subset are reported in **panel C**.

**A.**

| Setting | Type of SNL strategy | Type of SL pairs | PAGAN p2p Average AUROC | GNN edge prediction Average AUROC | SLGNN AUROC |
|---|---|---|---|---|---|
| A | Unmatched | All | 95.51±0.28 % | 93.27±2.12 % | 53.55% |
| B | Unmatched | CRISPR | 87.88±1.89 % | 94.62±0.40 % | 60.86% |
| C | Matched | All | 86.57±0.70 % | 83.12±4.50 % | 49.22% |
| D | Matched | CRISPR | 80.33±3.52 % | 85.34±2.00 % | 37.93% |

**B.**

| Setting | Type of SNL strategy | Type of SL pairs | PAGAN p2p Average AUPR | GNN edge prediction Average AUPR | SLGNN AUPR |
|---|---|---|---|---|---|
| A | Unmatched | All | 81.51±1.81 % | 73.20±2.92 % | 15.13% |
| B | Unmatched | CRISPR | 18.55±4.11 % | 34.81±2.05 % | 5.04% |
| C | Matched | All | 42.67±2.41 % | 38.12±3.97 % | 10.17% |
| D | Matched | CRISPR | 13.89±2.44 % | 16.96±2.44 % | 2.27% |

**C.**

| Setting | Type of SNL strategy | Type of SL pairs | Size of SL train set | Size of SL test set | Size of SNL train set | Size of SNL test set | Unique number of genes in the train set | Unique number of genes in the test set |
|---|---|---|---|---|---|---|---|---|
| A | Unmatched | All | 3386 | 199 | 3386 | 1825 | 6341 | 3135 |
| B | Unmatched | CRISPR | 966 | 75 | 966 | 3928 | 2085 | 6973 |
| C | Matched | All | 3386 | 199 | 3386 | 1825 | 3550 | 1798 |
| D | Matched | CRISPR | 966 | 90 | 966 | 2702 | 1181 | 791 |

**Supplementary Table 3. Test performance of the PAGAN approach in the pairs-to-pairs (p2p) setting for synthetic lethality prediction in human.** Test performances are reported for the classification of Synthetic Lethal (SL) versus Synthetic Non-Lethal (SNL) gene pairs using balanced test sets. The Area Under the Receiver Operating Characteristic curve (AUROC; **panel A**) and the Area Under the Precision–Recall curve (AUPR; **panel B**) are shown for different experimental settings defined by the strategy used to generate human SNL pairs through random sampling of non-essential genes. Settings **A** and **B** correspond to the unmatched strategy (i.e. in each SNL pair, one gene was sampled from the pool of non-essential genes involved in SL pairs and the other from non-essential genes not found in SL pairs), whereas settings **C** and **D** correspond to the matched strategy (i.e. both genes in each SNL pair were sampled exclusively from non-essential genes not found in SL pairs). In addition, settings **A** and **C** include SL pairs validated in vitro by either CRISPR or RNAi experiments, while settings **B** and **D** include only SL pairs supported by CRISPR experiments (**Methods**)**.** Results are presented for the PAGAN approach applied in the pairs-to-pairs setting (denoted PAGAN p2p) as well as for the corresponding Graph Neural Network (GNN) edge-prediction approach trained on identical data subsets. Performance values are reported as the mean and standard deviation across the six hyperparameter combinations evaluated. In addition, performances are also reported for SLGNN, a GNN–based method representative of the state-of-the-art methods for SL prediction in human, and considered here as a baseline. The total numbers of SL and SNL pairs included in the training and test sets for each subset are reported in **panel C**.

| DMF | Training setting | PAGAN g2p Average AUROC | PAGAN g2p Max AUROC | PAGAN g2p Average AUPR | PAGAN g2p Max AUPR |
|---|---|---|---|---|---|
| 0.5 | All individual genes | 66.68±2.45 % | 71.07% (32, 'relu') | 64.74±2.89 % | 69.47% (32, 'relu') |
| 0.4 | All individual genes | 66.90±2.77 % | 72.05% (32, 'relu') | 65.11±2.91 % | 70.08% (32, 'relu') |
| 0.3 | All individual genes | 69.36±2.61 % | 74.25% (32, 'relu') | 67.68±2.49 % | 71.97% (32, 'relu') |
| 0.5 | Essential gene-overlap controlled training | 66.19±2.21 % | 68.62% (32, 'relu') | 64.43±1.64 % | 66.60% (32, 'relu') |
| 0.4 | Essential gene-overlap controlled training | 66.36±2.31 % | 69.10% (32, 'relu') | 64.85±1.72 % | 67.26% (32, 'relu') |
| 0.3 | Essential gene-overlap controlled training | 68.67±2.72 % | 72.12% (32, 'relu') | 67.24±2.35 % | 70.28% (32, 'relu') |

**Supplementary Table 4. Test performance of the PAGAN approach in the genes-to-pairs (g2p) setting for synthetic lethality prediction in yeast by a model trained to classify yeast essential genes.** Test performances are reported for the classification of Synthetic Lethal (SL) versus Synthetic Non-Lethal (SNL) gene pairs achieved by the PAGAN approach applied in the genes-to-pairs setting (denoted PAGAN g2p). Two trainins settings were evaluated: in the first, denoted in the table as "All individual genes", PAGAN models were trained to classify the 1311 essential and 5268 non-essential genes in the knowledge-graph. In a second setting, denoted in the table as "Essential gene-overlap controlled training », the non-essential genes participating in SL or SNL pairs were excluded from the training set, i.e. minimizing the risk of data leakage between train and test sets under the genes-to-pairs setting. The Area Under the Receiver Operating Characteristic curve (AUROC) and the Area Under the Precision–Recall curve (AUPR) are shown for SL pairs defined using increasingly stringent thresholds of double-mutant fitness (DMF): DMF < 0.5, DMF < 0.4, and DMF < 0.3. Performance values are reported as the mean and standard deviation across the six hyperparameter combinations evaluated (third and fifth columns ; Methods). In addition, the maximum AUROC and AUPR values achieved by any individual hyperparameter configuration are provided (fourth and sixth columns; **Methods**), together with the corresponding hyperparameters indicated in parentheses, specifying the embedding dimension and activation function ('relu' or 'tanh').

**A.**

| Cell line | SL pairs in test set | PAGAN n2n training set | PAGAN g2p Average AUROC | PAGAN g2p Max AUROC | PAGAN g2p Average AUPR | PAGAN g2p Max AUPR |
|---|---|---|---|---|---|---|
| K562 | All | All individual genes | 49.72±1.21 % | 50.98% (64, | 37.06±0.90 % | 38.10% (64, 'tanh') |
| A375 | All | All individual genes | 69.16±6.22 % | 75.09% (64, | 46.43±5.50 % | 51.15% (64, 'tanh') |
| A549 | All | All individual genes | 79.74±1.37 % | 80.92% (32, 'relu') | 60.84±1.85 % | 62.23% (16, 'relu') |
| K562 | CRISPR | All individual genes | 81.15±0.73 % | 82.09% (32, 'relu') | 43.08±2.09 % | 45.32% (64, 'tanh') |
| A375 | CRISPR | All individual genes | 92.98±2.89 % | 96.76% (64, 'relu') | 2.53±1.10 % | 4.42% (64, 'relu') |
| A549 | CRISPR | All individual genes | 68.91±2.14 % | 71.48% (32, 'relu') | 7.23±0.51 % | 7.73% (64, 'relu') |
| K562 | All | Essential gene-overlap controlled | 49.78±1.82 % | 51.61% (16, 'relu') | 37.26±0.62 % | 38.27% (64, 'relu') |
| A375 | All | Essential gene-overlap controlled | 71.02±4.25 % | 76.59% (32, 'relu') | 47.55±4.67 % | 54.54% (64, 'tanh') |
| A549 | All | Essential gene-overlap controlled | 80.17±1.48 % | 82.26% (32, 'relu') | 62.19±3.02 % | 66.30% (32, 'relu') |
| K562 | CRISPR | Essential gene-overlap controlled | 81.37±0.88 % | 82.91% (64, 'relu') | 43.58±2.33 % | 46.24% (64, 'relu') |
| A375 | CRISPR | Essential gene-overlap controlled | 94.36±2.08 % | 97.24% (64, 'relu') | 3.17±1.38 % | 5.21% (64, 'relu') |
| A549 | CRISPR | Essential gene-overlap controlled | 70.23±1.18 % | 72.10% (32, 'relu') | 8.03±0.84 % | 9.15% (32, 'relu') |

**B.**

| Cell line | PAGAN genes-to-genes training set | # Essential genes | # Non essential genes | # SL pairs in test set (All) | # SL pairs in test set (CRISPR) | # SNL pairs in test set |
|---|---|---|---|---|---|---|
| K562 | All individual genes | 892 | 18438 | 5211 | 1267 | 10422 |
| A375 | All individual genes | 913 | 18417 | 938 | 5 | 1876 |
| A549 | All individual genes | 906 | 18424 | 831 | 75 | 1662 |
| K562 | Essential gene-overlap controlled | 892 | 7377 | 5211 | 1267 | 10422 |
| A375 | Essential gene-overlap controlled | 913 | 15080 | 938 | 5 | 1876 |
| A549 | Essential gene-overlap controlled | 906 | 16191 | 831 | 75 | 1662 |

**Supplementary Table 5. Test performance of the PAGAN approach in the genes-to-pairs setting for cell-line–specific synthetic lethality prediction in human. Panel A** reports the test performances on the classification of Synthetic Lethal (SL) versus Synthetic Non-Lethal (SNL) gene pairs achieved by the PAGAN approach applied in the genes-to-pairs setting. Performances are shown for three independent, cell-line–specific PAGAN models trained to classify genes according to cell-line–specific gene essentiality/non-essentiality, using corresponding essential and non-essential gene lists for the K562, A375, and A549 cell lines (Methods). Two settings for the PAGAN training were considered: one considering all essential and non-essential genes for a given cell line, and another one excluding from the training set the non-essential genes participating in SL or SNL pairs**.** For each cell line, PAGAN models were evaluated on their ability to discriminate the corresponding sets of SL and SNL gene pairs, considering either all experimentally validated in vitro SL pairs or only SL pairs established through CRISPR experiments, as indicated in the table (2$^{nd}$ column). The Area Under the Receiver Operating Characteristic curve (AUROC) and the Area Under the Precision–Recall curve (AUPR) are reported as the mean and standard deviation across the six hyperparameter combinations evaluated (fourth and sixth columns; Methods). In addition, the maximum AUROC and AUPR values achieved by any individual hyperparameter configuration are provided (fifth and seventh columns), together with the corresponding hyperparameters specified in parentheses, indicating the embedding dimension and activation function ('relu' or 'tanh'). **Panel B** reports the total numbers of essential and non-essential genes included in the training set, together with the numbers of SL and SNL gene pairs included in the test sets for each cell line and experimental condition.
